# "Unveiling Prostate Cancer’s Molecular Tapestry: Ethnic Disparities and Prognostic Insights from Whole-Mount Prostatectomy Tissue Analysis"

**DOI:** 10.1101/2024.07.15.24310431

**Authors:** Wei Zhao, Pin Li, Shannon Carskadon, Sunita Ghosh, Craig Rogers, James Peabody, Dhananjay Chitale, Mani Menon, Sean Williamson, Nilesh Gupta, Nallasivam Palanisamy

**Affiliations:** Department of Hematology/Oncology, Henry Ford Health, Detroit, MI, USA; Department of Public Health, Henry Ford Health, Detroit, MI, USA; Department of Urology, Henry Ford Health, Detroit, MI, USA; Department of Pathology, Henry Ford Heath, Detroit, MI, USA; Department of Biomaterials, Saveetha Dental College, and Hospitals, Saveetha Institute of Medical and Technical Sciences, Saveetha University, Chennai, India

**Keywords:** Prostate cancer, tumor heterogeneity, molecular biomarkers, gene fusion

## Abstract

**Background:** Prostate cancer is a complex and heterogeneous disease with multiple tumor foci, each potentially harboring distinct driver molecular aberrations. This complexity poses challenges to effective management. We took an innovative approach to gain a comprehensive understanding of the genetic underpinnings of each tumor focus and avoid overlooking more minor yet clinically significant foci. Instead of relying solely on a systematic sampling of dominant foci, we conducted molecular analysis on whole-mount radical prostatectomy specimens. Our study aimed to find distinct molecular subsets of prostate cancer and assess their correlation with clinical outcomes, focusing on Caucasians (CA) and African Americans (AA).

**Method:** We randomly selected 2201 whole-mount radical prostatectomy cases, with 1207 (54.8%) from CA and 994 (45.1%) from AA patients evaluated for a 5-year biochemical recurrence-free survival rate (BCR). Of these 2201 cases, 834 (463 −56% were from CA and 371 −44% from AA patients) were subjected to molecular analysis using dual immunohistochemistry (IHC) for ERG and SPINK1, along with dual RNA *in-situ* hybridization (RNA-ISH) for ETV1 and ETV4 to evaluate tumor molecular heterogeneity on whole-mount specimens. The Chi-squared test examined racial disparities in aberrant oncogene expression. To assess BCR-free survival, we employed the Kaplan-Meier method and Cox-PH models for patients with distinct molecular subsets of prostate cancer. Additionally, Gleason Grade groups of prostate biopsies were summarized using a spaghetti plot and compared using linear mixed models.

**Results:** Analysis of the 2201 cases revealed that AA with localized prostate cancer behaved differently with better 5-year BCR-free survival than CA after radical prostatectomy (AA: 0.82, 95% CI 0.80-0.85; CA: 0.71, 95% CI 0.68-0.75; p<.001). Molecular profiling of whole-mount specimens from 834 cases revealed that 16.4%, 58.4%, 21.7%, and 3.5% of patients with localized prostate cancer expressed none, one, two, and three of the four oncogenes, respectively. This finding identified new molecular subsets of prostate cancer with more than one driver mutation in a mutually exclusive manner within the multifocal disease. ERG and SPINK1 expression showed a negative correlation (p<.001). Notably, AA patients exhibited a lower incidence of ERG (38.8% vs. 60.3%) but a higher incidence of SPINK1 (63.3% vs. 35.6%) than CA patients. The incidences of ETV1 (9.4% vs. 9.3%) and ETV4 (4.6% vs. 3.9%) were not statistically significant between the two racial groups. However, significantly, ETV1 expression was associated with worse BCR-free survival in CA patients (hazard ratio [HR]=2.36, 95% CI 1.22-4.57, p=.02), while ETV4 expression was linked to worse BCR-free survival in AA patients (HR=2.65, 95% CI 1.15-6.09, p=.02). Moreover, ETV4 expression was associated with regional lymph node metastasis in AA patients (odds ratio [OR]=5.14, 95% CI 1.3-17.4, p=.01) but not in CA patients (OR=0.60, 95% CI 0.03-3.17, p=.63) at the time of radical prostatectomy. Additionally, in patients who underwent multiple biopsies before radical prostatectomy, the Gleason Grade group increased over time in AA patients (0.25 per year, p<.001) but remained unchanged in CA patients. ERG expression was associated with a lower Gleason Grade group (−0.20, p=.03), while ETV4 expression was linked to a higher Gleason Grade (0.54, p=.01).

**Conclusions:** Our study reveals that AA with localized prostate cancer behaves differently and has better BCR-free survival than CA after radical prostatectomy, even after adjusting for known prognostic factors. Identification of new molecular subsets of prostate cancer with more than one ETS gene fusion within a multifocal prostate shows significant molecular heterogeneity between localized prostate cancer in CA and AA patients. Importantly, given the association of ETV1 and ETV4 expression with worse BCR-free survival in CA and AA, respectively, ETV1 and ETV4 emerge as potential prognostic markers, offering insights for clinical practice to predict prostate cancer recurrence after radical prostatectomy. Identification of new molecular subsets of prostate cancer with more than one ETS gene fusion and SPINK1 in a mutually exclusive pattern indicates the clonal origin of independent tumor foci, which is a rare and unique phenomenon in prostate cancer hitherto unidentified.

## 1. INTRODUCTION

Prostate cancer is a complex disease characterized by distinct morphological heterogeneity in a multifocal disease. The genetic underpinnings of each tumor focus have yet to be studied extensively. Each tumor focus is thought to have unique driver mutations. Understanding the extent of tumor molecular heterogeneity will allow us to categorize prostate cancer into various molecular subtypes. A prevalent genomic alteration in this context is the fusion of E26 transformation-specific (ETS) family transcription factors with androgen-regulated, prostate-specific genes^1^. These rearrangements result in overexpression that is not typically found in benign prostate tissue. Among the ETS gene fusions, ERG is the most often occurring in prostate cancer, followed by SPINK1, ETV1, and ETV4. Although ERG, ETV1, and ETV4 belong to the ETS family transcription factors, they have an independent role in prostate cancer. We have reported the independent clonal evolution of tumors with distinct driver molecular aberrations for these markers using needle biopsy and whole-mount prostatectomy specimens^2, 3^. It has been shown that ERG and ETV1 control a common transcriptional network but in opposing fashions. ERG negatively regulates the androgen receptor (AR) transcriptional program, while ETV1 cooperates with AR signaling by favoring the activation of the AR transcriptional program. Unlike ERG, ETV1 expression promotes autonomous testosterone production and up-regulates the expression of AR target genes and genes involved in steroid biosynthesis and metabolism. ETV1 also supports the development of invasive adenocarcinoma under the background of complete PTEN loss. The distinct biology of ETV1-associated prostate cancer suggests that this disease class may need new therapies directed to underlying programs controlled by ETV1^4^. It has been shown that ERG and ETV1 have shared and distinct chromatin targets, influencing AR signaling differently. Notably, ERG expression is enriched in localized prostate cancer, while ETV1 expression is more common in high Gleason grade and metastatic prostate cancer, as supported by two independent cohorts^4^. Additionally, ETV4 promotes metastasis in response to the activation of PI3-kinase and Ras signaling in an advanced prostate cancer mouse model^5, 6, 7^.

Prostate cancer cases lacking ETS fusions may show malignancy due to abnormal expression of serine peptidase inhibitor Kazal type 1 (SPINK1), which activates the epithelial growth factor receptor (EGFR) because of its structural similarity to epithelial growth factors (EGF). SPINK1, commonly known for its role in pancreatic function, is also associated with aggressive prostate cancer subtypes, including those resistant to androgen deprivation therapy. SPINK1 is more widely expressed in African American (AA) prostate cancer patients than in Caucasian American (CA) patients^2, 3, 8^. However, the relationship between SPINK1 expression and biochemical recurrence (BCR) after radical prostatectomy is still uncertain, given conflicting evidence. Recent findings show that SPINK1-positive tumors may have a distinct molecular profile, potentially influencing disease aggressiveness and therapeutic response^9^.

SPINK1-positive prostate cancer is linked to high-grade tumors and an increased likelihood of lymph node and distant metastasis. The expression of SPINK1 disrupts the AR signaling pathway, promoting cancer cell invasion and migration. Elevated SPINK1 levels in tumor tissues or blood samples are a potential biomarker for lymph node involvement, suggesting potential clinical significance in prognosis and disease management. Targeting SPINK1 may offer strategies for treating this subtype of prostate cancer^10, 11^. We showed the molecular mechanism of SPINK1 overexpression mediated by the epigenetic silencing of miRNA 338-5p and miRNA-421^12^, and androgen deprivation upregulates SPINK1 expression and potentiates cellular plasticity in prostate cancer. Using an integrated proteomics approach, we showed tyrosine kinase KIT as a therapeutic target for SPINK1-positive prostate cancer^13^. These findings underscore the importance of understanding the expression pattern in multifocal cancer and its impact on clinical outcomes. Such insights may pave the way for targeted therapies tailored to the specific molecular subtypes of prostate cancer, improving treatment outcomes and patient prognosis.

Prostate cancer presents striking racial disparities, with AA facing a twofold higher mortality rate than CA. Socioeconomic barriers contribute to this outcome by hindering access to prompt screenings, early detection, and standard treatments. However, evidence suggests that AA can achieve improved prostate cancer-specific survival rates comparable to CA in equitable health systems or clinical trials^14^. Recent studies have further explored the implications of ETS gene fusions and SPINK1 in racial disparities^15^. For instance, investigations have shown a higher prevalence of ERG fusions in CA prostate cancer patients compared to AA, suggesting a possible molecular basis for differential disease progression and response to treatment^9, 15^. Moreover, emerging research has identified additional molecular alterations, such as SPOP mutations and FOXA1 alterations, that exhibit distinct racial patterns and may contribute to the observed disparities in prostate cancer outcomes^16^. Despite the well-known oncogenic potential of these molecular alterations in prostate cancer, their ability to reliably predict patient prognosis remains limited. This discrepancy may stem from the sampling methods used in the earlier studies, which often focused on the dominant tumor focus while neglecting smaller foci with potential clinical significance. Moreover, correlations of driver mutations with clinical outcomes were typically studied in mixed populations without acknowledging racial differences in tumor genomics between AA and CA patients. To address these limitations, we comprehensively analyzed ERG, ETV1, ETV4, and SPINK1 expression in whole-mount radical prostatectomy specimens from AA and CA patients with localized prostate cancer. By adopting a race-sensitive approach, we correlated the expression of these driver mutations with clinical outcomes. By considering racial disparities and employing a robust sample collection method, our study aimed to advance our understanding of the prognostic implications of these molecular alterations in prostate cancer.

## 2. METHODS

### 2.1 Study design

We randomly selected and reviewed clinical data for 1207 (54.8%) CA and 994 (45.1%) AA patients with localized prostate cancer and evaluated their 5-year biochemical recurrence (BCR)-free survival rate. Of the 2201 cases, we collected whole-mount radical prostatectomy specimens for 834 cases from the Department of Pathology and Laboratory Medicine archives at Henry Ford Health.

These specimens were obtained from patients who underwent radical prostatectomy for localized prostate cancer before any treatment between April 2001 and June 2018. Comprehensive clinical data, including their age, race, family history of cancer, Gleason Grade groups from biopsies taken before prostatectomy, preoperative and postoperative prostate-specific antigen (PSA) levels, and BCR status were collected. BCR was defined as an increase in PSA level from undetectable to 0.2 ng/mL or above postoperatively and confirmed by a second measurement. The total number of biopsies before radical prostatectomy was recorded. The pathological features of each whole-mount radical prostatectomy specimen were thoroughly examined, including information such as tumor location, tumor volume, dominant focus, secondary foci, Gleason Grade group, morphology, pathological T stage (pT), margin status, and lymph node status. Each specimen’s representative whole-mount tissue section having the dominant/index tumor foci was chosen to further evaluate selected markers through immunohistochemistry (IHC) and RNA *in situ* hybridization (RNA-ISH). This comprehensive data collection and analysis aim to shed light on the molecular characteristics of each tumor focus and clinical outcomes of patients who underwent radical prostatectomy, providing significant insights into the behavior of localized prostate cancer and its potential for disease progression and recurrence.

### 2.2 Histological evaluation

Tumor location (posterior aspect vs. anterior aspect) was assessed, and tumor volume (≤5%, 6%-10%, 11%-20%, and >20%) was estimated from whole-mount slides by pathologists specializing in genitourinary cancer. The estimation of tumor volume combined all foci accumulated across the entire gland. The dominant focus was designated if it carried the most pathological significance, considering factors such as the highest Gleason score, the largest tumor size if the Gleason scores were similar among foci, the shortest distance from resection margins, and positive resection margins. The less pathologically significant tumor foci were designated as secondary, including smaller tumor sizes, irrespective of Gleason scores or locations further from resection margins. We used a minimum distance of 3 mm to distinguish anatomically independent tumor foci. One representative whole-mount prostate tissue section was chosen according to the following criteria: the section included the dominant focus with the most significant tumor volume, or both dominant and secondary foci if multiple foci were present, and/or both dominant focus and clinically significant secondary foci, defined as those with Gleason Grade Group 4 features. We also recorded the number of total tumor foci, Gleason Grade group, pathological T stage, resection margin status, and lymph node status.

### 2.3 Dual IHC for ERG and SPINK1 and dual RNA-ISH for ETV1 and ETV4

We have developed methods for simultaneously evaluating the selected four markers by dual IHC and dual RNAISH methods^3, 17, 18^. IHC was used to detect ERG and SPINK1 protein expression, while RNA-ISH (RNAscope) was selected to detect ETV1 and ETV4 mRNA expression due to the unavailability of cancer-specific monoclonal antibodies^2^. For RNA-ISH, the whole-mount slides were initially incubated at 60°C for 1 hour, then deparaffinized using xylene twice for 5 minutes each with periodic agitation. The slides were then immersed in 100% ethanol twice for 3 minutes, with occasional agitation, and air-dried for 5 minutes. After encircling the tissues with a pap pen and treating them with H2O2 for 10 minutes, the slides were rinsed in distilled water and boiled in 1X Target Retrieval for 15 minutes. Subsequently, they were treated with Protease Plus for 15 minutes at 40°C in a HybEZ Oven (Bio-Techne, 310010). These steps were facilitated by the RNAscope Pretreatment kit (Bio-Techne, 310020). Following further rinses in distilled water, the slides were exposed to ETV1 (Bio-Techne, 311411) and ETV4 (Bio-Techne, 478571-C2) probes at a 50:1 ratio for 2 hours at 40°C in the HybEZ Oven. Two washes in 1X Wash Buffer (Bio-Techne, 310091) for 2 minutes each were performed, and the slides were stored overnight in a 5X SSC solution. The slides were washed twice in 1X Wash Buffer for 2 minutes each the next day. Then, they underwent treatment with Amp 1, Amp 2, Amp 3, and Amp 4 for specific durations at 40⁰C in the HybEZ oven, with two washes in 1X Wash Buffer for 2 minutes each after each step. Subsequently, the slides were treated with Amp 5 and Amp 6 for specified durations at room temperature in a humidity chamber, again with two washes in 1X Wash Buffer for 2 minutes each after each step. Red color development was achieved by adding a 1:60 solution of Fast Red B: Fast Red A to each slide and incubating for 10 minutes. The slides were washed twice in 1X Wash Buffer for 2 minutes each, then treated with Amp 7 and Amp 8 at specified durations at 40⁰C in the HybEZ oven, with two washes in 1X Wash Buffer for 2 minutes each after each step. Further treatment with Amp 9 and Amp 10 was performed for specified durations at room temperature in a humidity chamber, followed by two washes in 1X Wash Buffer for 2 minutes each after each step. Brown color development (ETV1) was achieved by adding a solution of Betazoid DAB (1 drop DAB to 1ml Buffer: Biocare Medical, BDB2004) to each slide and incubating for 10 minutes. Amps 1-10 and Fast Red were (ETV4) included in the RNAscope 2.5 HD Duplex Detection Reagents (Bio-Techne, 322500). The slides were then washed twice in distilled water, followed by treatment with EnVision FLEX Hematoxylin (DAKO, K8008) for 5 minutes, rinsed in tap water several times, and dried completely. The slides were then dipped in xylene approximately 15 times, and EcoMount (Biocare Medical, EM897L) was added to each slide, which was then cover-slipped.

For IHC, slides were baked for 2 hours at 60°C, then placed in EnVision FLEX Target Retrieval Solution, high pH (Agilent DAKO, K800421-2) in a PT Link instrument (Agilent DAKO, PT200) at 75⁰C, heated to 97⁰C for 20 minutes, and then cooled to 75⁰C. Slides were washed in 1X EnVision FLEX Wash Buffer (DAKO, K8007) for 5 minutes. Subsequently, the slides were treated with Peroxidazed 1 (Biocare Medical, PX968M) for 5 minutes and Background Punisher (Biocare Medical, BP974L) for 10 minutes, with a wash of 1X EnVision FLEX Wash Buffer for 5 minutes after each step. Anti-ERG (EPR3864) rabbit monoclonal primary antibody (1:50; Abcam, ab92513) and a mouse monoclonal against SPINK1 (1:100; Novus Biologicals, H00006690-M01) were added to each slide, which was then cover-slipped with parafilm, placed in a humidifying chamber, and incubated overnight at 4⁰C. The following day, the slides were washed in 1X EnVision Wash Buffer for 5 minutes and then set in Mach2 Doublestain 1 (Biocare Medical, MRCT523) for 30 minutes at room temperature in a humidifying chamber. After rinsing the slides in 1X EnVision Wash Buffer 3 times for 5 minutes each, Warp Red solution (1 drop to 2.5ml buffer; Biocare Medical, WR806H) was added, and slides were incubated for 5 minutes. Alternatively, Ferangi Blue solution (1 drop to 2.5ml buffer; Biocare Medical, FB813) can be used. Then, slides were washed in 1X EnVision FLEX Wash Buffer for 5 minutes and treated with a Betazoid DAB solution (1 drop to 1ml buffer; Biocare Medical, BDB2004) for 5 minutes. The slides were then rinsed twice in distilled water, followed by treatment with EnVision FLEX Hematoxylin (DAKO, K8008) for 5 minutes. After several rinses in tap water, the slides were dried completely. Finally, the slides were dipped in xylene approximately 15 times, and EcoMount was added to each slide, which was then cover-slipped.

### 2.4 Use of RNA-ISH rather than IHC for ETV1 and ETV4

We utilized RNA-ISH to examine the expression of ETV1 and ETV4 mRNA and IHC to examine the expression of ERG and SPINK1 protein in radical prostatectomy specimens. We opted for an RNA-based technique over a protein-based one like IHC due to the absence of specific monoclonal antibodies. This is a viable alternate approach for markers lacking specific antibodies. Further, we have tested the specificity of each RNA-ISH probe for cross-reactivity with other ETS genes on positive control specimens with chromosome rearrangement in ERG, ETV1, ETV4, and ETV5 genes validated by FISH^18^.

### 2.5 Statistical analysis

The clinical and pathological characteristics of the study population were presented using median and interquartile range (IQR) for continuous data and counts and percentages for categorical data. The chi-square test was employed to compare the expression status of ERG, ETV1, ETV4, or SPINK1, and other categorical variables in whole-mount radical prostatectomy specimens between AA and CA with prostate cancer. Wilcoxon rank test was used to compare the continuous variables between AA and CA. BCR-free survival was defined as the time interval between radical prostatectomy and BCR, with censoring at the time of the last recorded PSA result. Within each racial group, BCR-free survival was compared between patients with positive and negative expression of these oncogenes using Kaplan-Meier curves with the log-rank test. Additionally, Cox-PH models were used to explore the correlation between the expression status of these oncogenes and BCR-free survival while adjusting for various prognostic factors, such as preoperative PSA levels, Gleason Grade group, pT, location of the primary focus, and histological subtypes. The correlation between the expression status (positive vs. negative) of these oncogenes and lymph node metastasis was analyzed using the chi-square test and odds ratios (OR), and the corresponding 95% confidence intervals (CI) were reported. The Gleason Grade groups of multiple biopsies taken before radical prostatectomy were visualized using a spaghetti plot. Linear mixed models were used to examine changes in Gleason Grade groups over time while considering race and oncogene expression status. Age at the first biopsy was included as a covariate. All statistical tests were two-sided, and a significance level of 0.05 (α) was used to determine statistical significance. R software version 4.0.4 (R Core Team, 2020) was used for statistical analysis.

## 3. RESULTS

### 3.1 Racial disparities in clinical outcomes of AA and CA with localized prostate cancer

In this study, we investigated the 5-year BCR-free survival in a large cohort of AA and CA patients who underwent radical prostatectomy for localized prostate cancer. We analyzed data from 1207 CA and 994 AA cases and observed racial differences in clinical outcomes. Interestingly, the study found that AA patients had a better prognosis compared to CA patients after adjusting for various prognostic factors (AA: 0.82, 95% CI 0.80-0.85; CA: 0.71, 95% CI 0.68-0.75, log-rank p<.001) (**Figure 1A**). After adjusting for various prognostic factors, race in CA remained to be significantly associated with worse 5-year BCR-free survival (Hazard Ratio [HR]=1.25, 95%CI 1.01-1.55) (**Figure 1B**). This suggests that there may be important racial disparities in the outcomes of patients with localized prostate cancer patients undergoing radical prostatectomy. Considering that socioeconomic and clinicopathological factors may not fully elucidate the observed racial disparities, we propose that variations in tumor genomics could potentially drive distinct tumor behaviors and clinical outcomes between AA and CA with localized prostate cancer.

**Figure 1:**
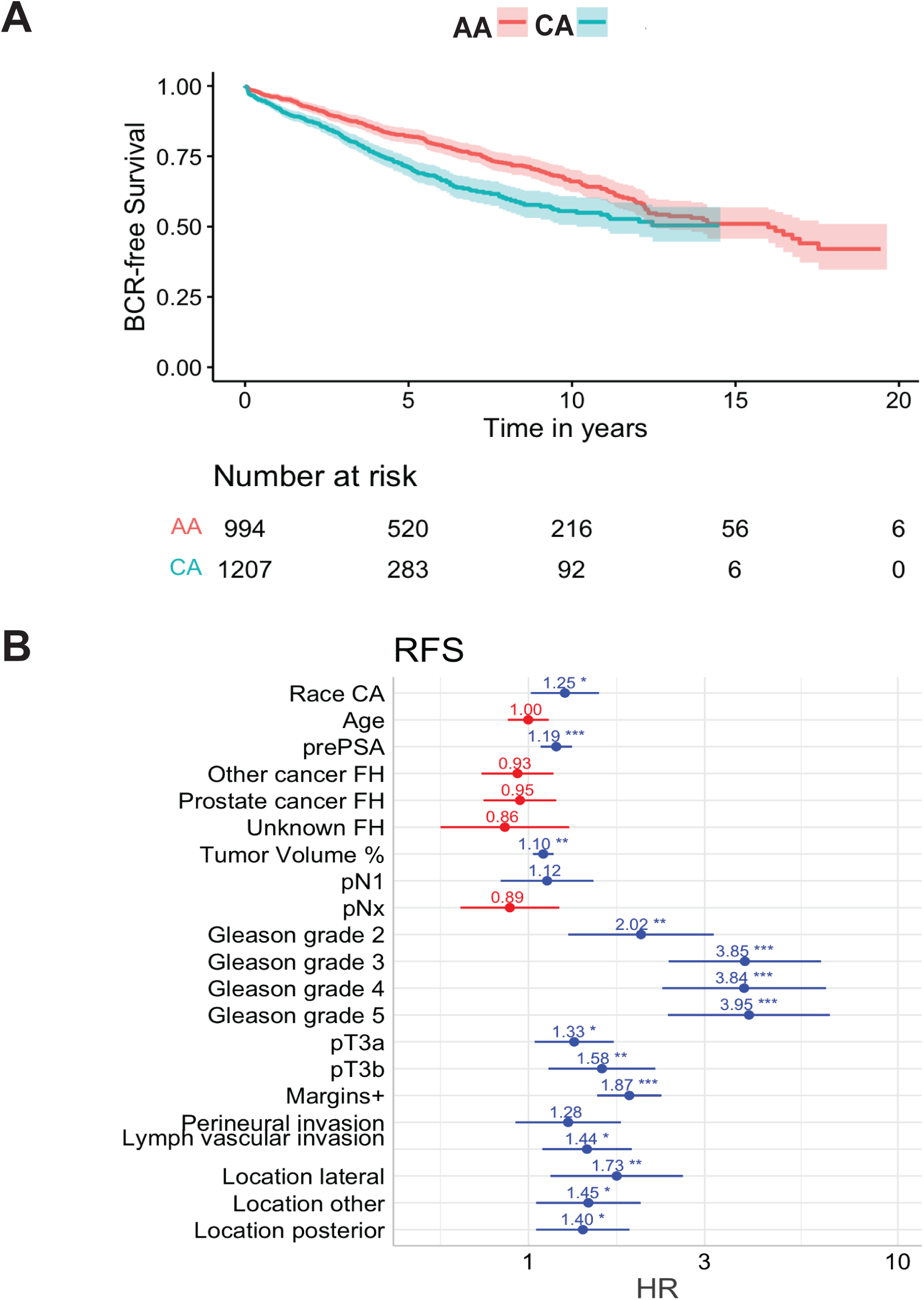
A) Biochemical recurrence free survival analysis show that prostate cancer in African Americans (AA) behaves differently with better survival than Caucasian American (CA) which show poor survival even after adjusting for known non-biological prognostic factors. B) Multivariate analysis for racial disparities in clinical outcomes between CA and AA.

### 3.2 Clinical and pathological characteristics of AA and CA with localized prostate cancer

Of the 2201 cases studied for BCR, we randomly selected a cohort of 834 patients who underwent radical prostatectomy for localized prostate cancer molecular profiling analysis. **Table 1** summarizes their clinical and pathological characteristics. Of the 834 cases, 371 (44%) were AA, while 463 (56%) were CA. Unlike conventional tissue microarray analysis using sampling of dominant/index tumor foci, this is the largest whole-mount radical prostatectomy cohort subjected to unbiased molecular profiling, including a large cohort of AA cases. The median age at the time of radical prostatectomy was 61, ranging from 34 to 83 years. AA patients exhibited significant differences from CA on various clinical and pathological parameters. Notably, AA had a higher incidence of a family history of prostate cancer (34.8% vs. 30.0%), a higher median preoperative PSA level (5.8 ng/ml vs. 5.4 ng/ml), and a higher prevalence of perineural invasion (51.8% vs. 35.9%). They also had a lower prevalence of lymphovascular invasion (8.4% vs. 14.7%) and a lower frequency of positive surgical margins (29.9% vs. 36.7%).

**Table 1.** Clinical and Pathological Characteristics of the Study Cohort.

|  |  | **African American** | **Caucasian** | **Total** | **p\*** |
| --- | --- | --- | --- | --- | --- |
| **Total N (%)** |  | 371 (44.5) | 463 (55.5) | 834 |  |
| **Age (years)** | Median (IQR) | 61.0 (55.0 to 67.0) | 62.0 (56.0 to 67.0) | 61.0 (56.0 to 67.0) | 0.203 |
| **Preoperative PSA (ng/mL)** | Median (IQR) | 5.8 (4.6 to 8.7) | 5.4 (4.2 to 7.6) | 5.6 (4.4 to 8.1) | 0.001 |
| **Family history** | Negative | 134 (36.1) | 134 (28.9) | 268 (32.1) | 0.001 |
|  | Other cancer | 93 (25.1) | 147 (31.7) | 240 (28.8) |  |
|  | Prostate cancer | 129 (34.8) | 139 (30.0) | 268 (32.1) |  |
|  | Unknown | 15 (4.0) | 43 (9.3) | 58 (7.0) |  |
| **Tumor volume (%)** | Median (IQR) | 10.0 (5.0 to 16.0) | 9.0 (5.0 to 17.0) | 10.0 (5.0 to 17.0) | 0.238 |
| **Tumor stage** | pT2 | 205 (55.6) | 261 (56.5) | 466 (56.1) | 0.592 |
|  | pT3a | 122 (33.1) | 138 (29.9) | 260 (31.3) |  |
|  | pT3b | 42 (11.4) | 63 (13.6) | 105 (12.6) |  |
| **Node stage** | pN0 | 299 (80.6) | 345 (74.5) | 644 (77.2) | 0.099 |
|  | pN1 | 26 (7.0) | 38 (8.2) | 64 (7.7) |  |
|  | pNx | 46 (12.4) | 80 (17.3) | 126 (15.1) |  |
| **Gleason Grade Group** | 1 | 39 (10.6) | 63 (13.7) | 102 (12.3) | 0.177 |
|  | 2 | 201 (54.8) | 225 (49.0) | 426 (51.6) |  |
|  | 3 | 92 (25.1) | 108 (23.5) | 200 (24.2) |  |
|  | 4 | 11 (3.0) | 17 (3.7) | 28 (3.4) |  |
|  | 5 | 24 (6.5) | 46 (10.0) | 70 (8.5) |  |
| **Tumor margins** | Negative | 260 (70.1) | 293 (63.3) | 553 (66.3) | 0.047 |
|  | Positive | 111 (29.9) | 170 (36.7) | 281 (33.7) |  |
| **Perineural invasion** | Absent | 179 (48.2) | 297 (64.1) | 476 (57.1) | <0.001 |
|  | Present | 192 (51.8) | 166 (35.9) | 358 (42.9) |  |
| **Lymph vascular invasion** | Absent | 340 (91.6) | 395 (85.3) | 735 (88.1) | 0.007 |
|  | Present | 31 (8.4) | 68 (14.7) | 99 (11.9) |  |
| **Time biopsy to surgery (months)** | Median (IQR) | 2.7 (1.8 to 4.2) | 2.5 (1.8 to 4.0) | 2.6 (1.8 to 4.1) | 0.208 |
| **Tumor location** | Anterior/anterolateral | 89 (24.0) | 104 (22.5) | 193 (23.1) | 0.226 |
|  | Lateral | 31 (8.4) | 51 (11.0) | 82 (9.8) |  |
|  | Other | 56 (15.1) | 52 (11.2) | 108 (12.9) |  |
|  | Posterior/posterolateral | 195 (52.6) | 256 (55.3) | 451 (54.1) |  |

\* Chi-squared test for categorical variables, Wilcoxon rank test for continuous variables. Abbreviations: IQR, interquartile range; PSA, prostate-specific antigen.

### 3.3 Molecular heterogeneity between AA and CA with localized prostate cancer

Our study examined the expression of ERG, ETV1, ETV4, and SPINK1 in whole-mount radical prostatectomy specimens. **Figures 2 & S2-S8** illustrate examples of the multifocal nature of prostate cancer, where separate tumor foci are surrounded by benign prostate tissue. Interestingly, the expression patterns of the four oncogenes were mutually exclusive within a single tumor focus but could coexist in different tumor foci within the same sample. We did not observe any tumor foci positive with more than one gene. No case showed the presence of a single marker in all the tumor foci, indicating the mutually exclusive expression patterns and independent clonal evolution of each tumor focus with or without ETS gene fusions and SPINK1. Among patients with localized prostate cancer, 16.4% expressed none of the four oncogenes, 58.4% expressed one, 21.7% expressed two, and 3.5% expressed three oncogenes. We compared patients’ clinical and pathological characteristics with and without the expression of individual oncogenes (**Table S1-S4**). ERG expression was more common in CA than in AA (60.3% vs. 38.8%). Conversely, SPINK1 expression was more common in AA than in CA (63.3% vs. 35.6%). Although there were differences in the expression of ETV1 or ETV4 between AA and CA, they were not statistically different (**Figure 3A**). Moreover, we observed a negative correlation between ERG and SPINK1 expression in AA and CA (AA: phi coefficient −0.13, p=.01; CA: phi coefficient −0.23, p<.001). In AA, we found a positive correlation between ERG, ETV1, and ETV4 expression (phi coefficient 0.10, p=.05; 0.12, p=.02) (**Figure 3B**). Based on the expression patterns of these four oncogenes, we identified new molecular subtypes of prostate cancer in both AA and CA (**Table 2**). We observed that the prevalence of ERG-positive, ETV1/EVT4/SPINK1-negative prostate cancer was lower in AA compared to CA (13.7% vs. 40.4%, p<.001). Conversely, the prevalence of SPINK1-positive, ERG/ETV1/ETV4-negative prostate cancer was higher in AA than in CA (38.3% vs. 17.3%, p<.001). These findings suggest the presence of distinct molecular subtypes of prostate cancer between the two racial groups, a discovery made possible only through unbiased analysis using whole-mount specimens.

**Figure 2:**
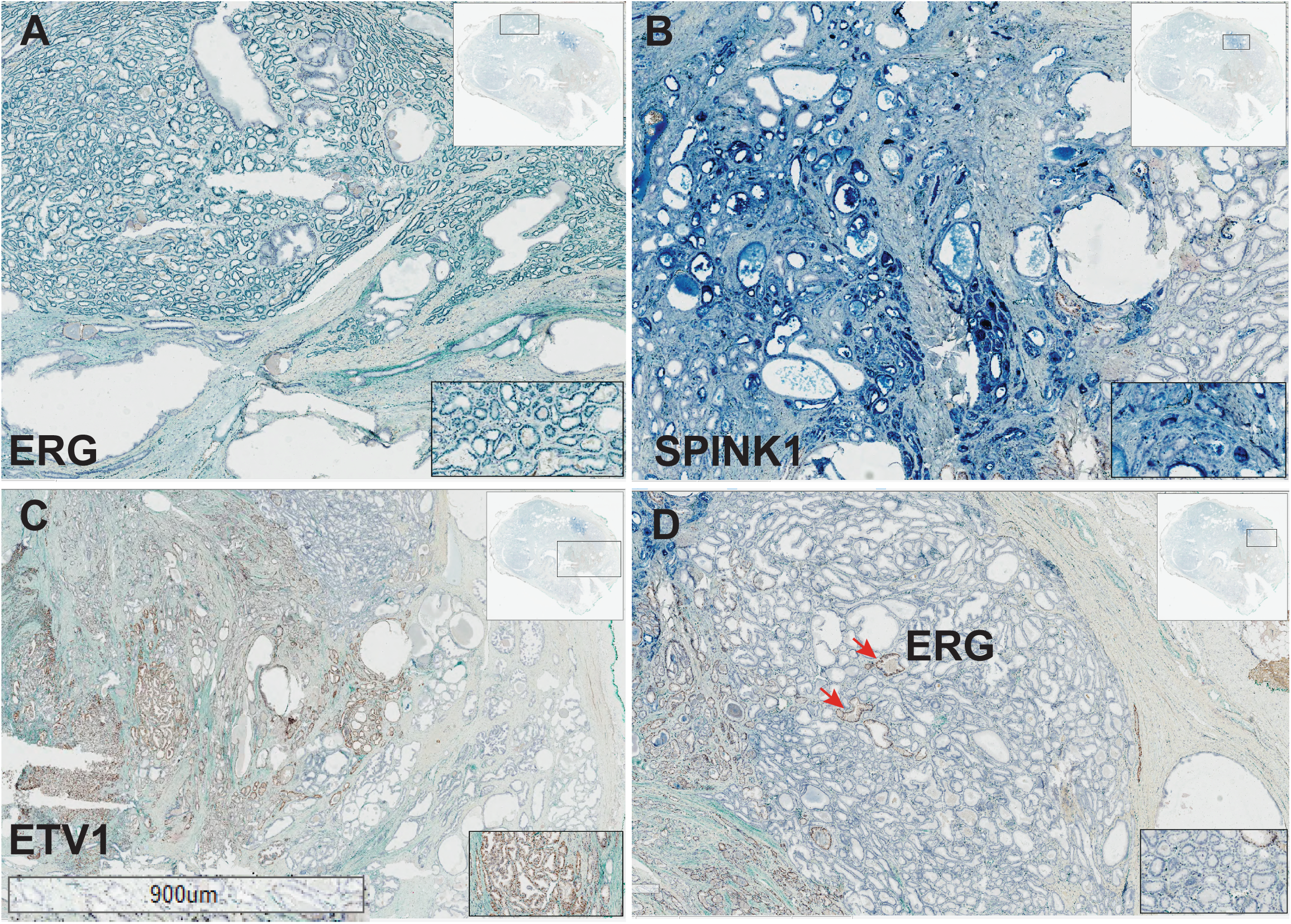
Wholemount radical prostatectomy tissue from a Caucasian American patient with multiple tumor foci with Gleason score 7 with tertiary Gleason pattern 5 with extraprostatic extension into left posterolateral region, pT3a. Gleason 3=83%, Gleason 4=15%, Gleason 5=2%, combined Gleason grade 7. Additional nodules at left posterolateral region. Simultaneous evaluation of ERG, SPINK1, ETV1 and ETV4 on a wholemount tissue with multifocal cancer (with five tumor foci) from a Caucasian American patient. Three of the five tumor foci were positive for ERG (A), SPINK1(B) and ETV1(C) and the other two foci were negative (D) for all four markers showing a mutually exclusive expression pattern indicating independent clonal origin of each tumor foci with distinct driver molecular aberrations. Arrows indicate small tumor foci positive for ERG within a large tumor foci showing the extent of intra tumor heterogeneity.Two additional tumor foci that were negative for all the four markers tested, potentially harbor unknown molecular aberration. Inset at the top right in each image show the wholemount view with the location of the tumor positive for a marker shown in rectangle. box. The inset at the bottom right show the zoom in view of the tumor foci.

**Figure 3:**
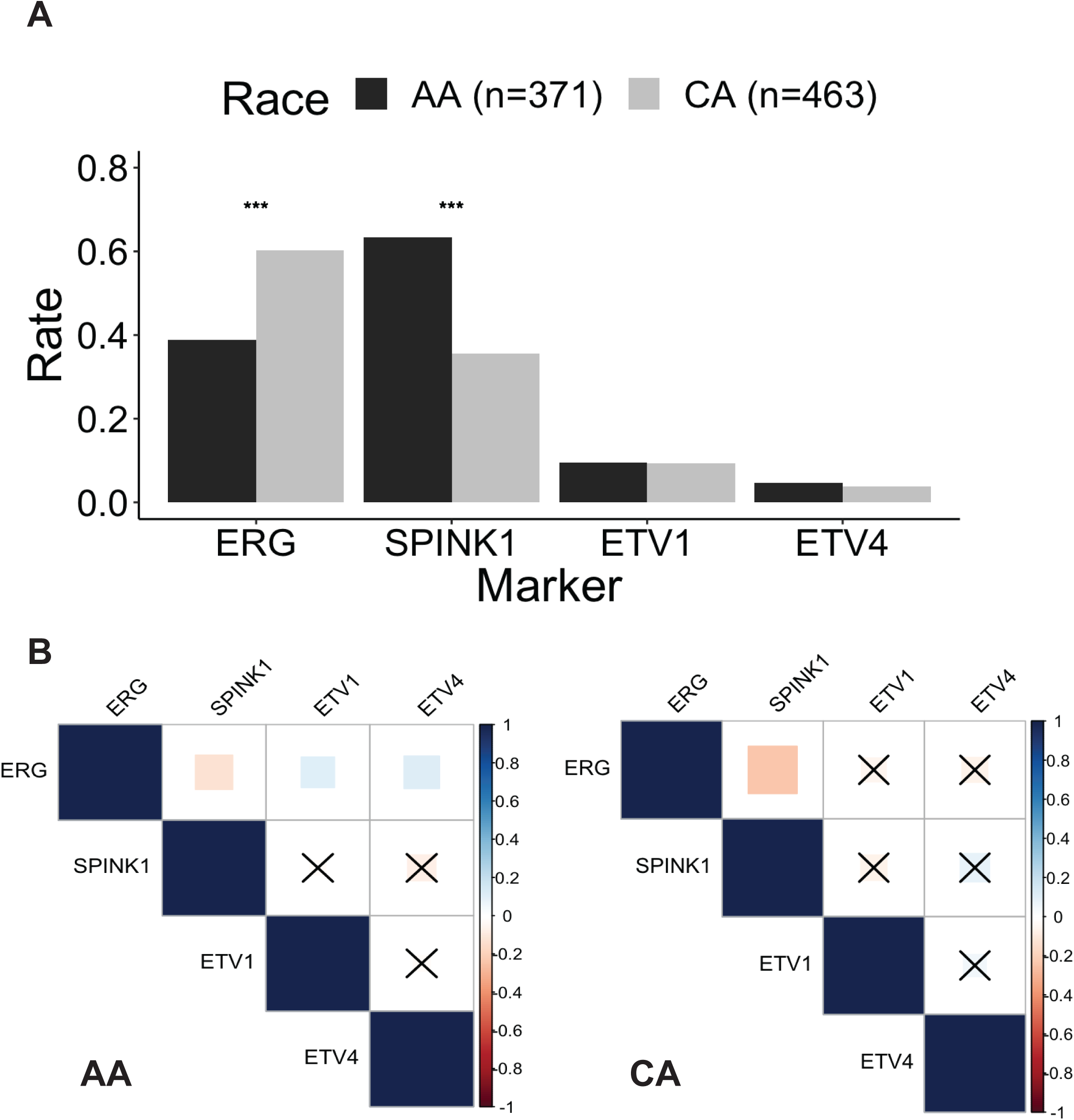
A) Bar plot comparing expression rates of ERG, SPINK1, ETV1, and ETV4 in prostate cancer between African Americans and Caucasian Americans. ***p<.001. (B) Heatmap showing the correlation between ERG, SPINK1, ETV1, and ETV4 expression in prostate cancer in African Americans and Caucasian Americans.

**Table 2.**
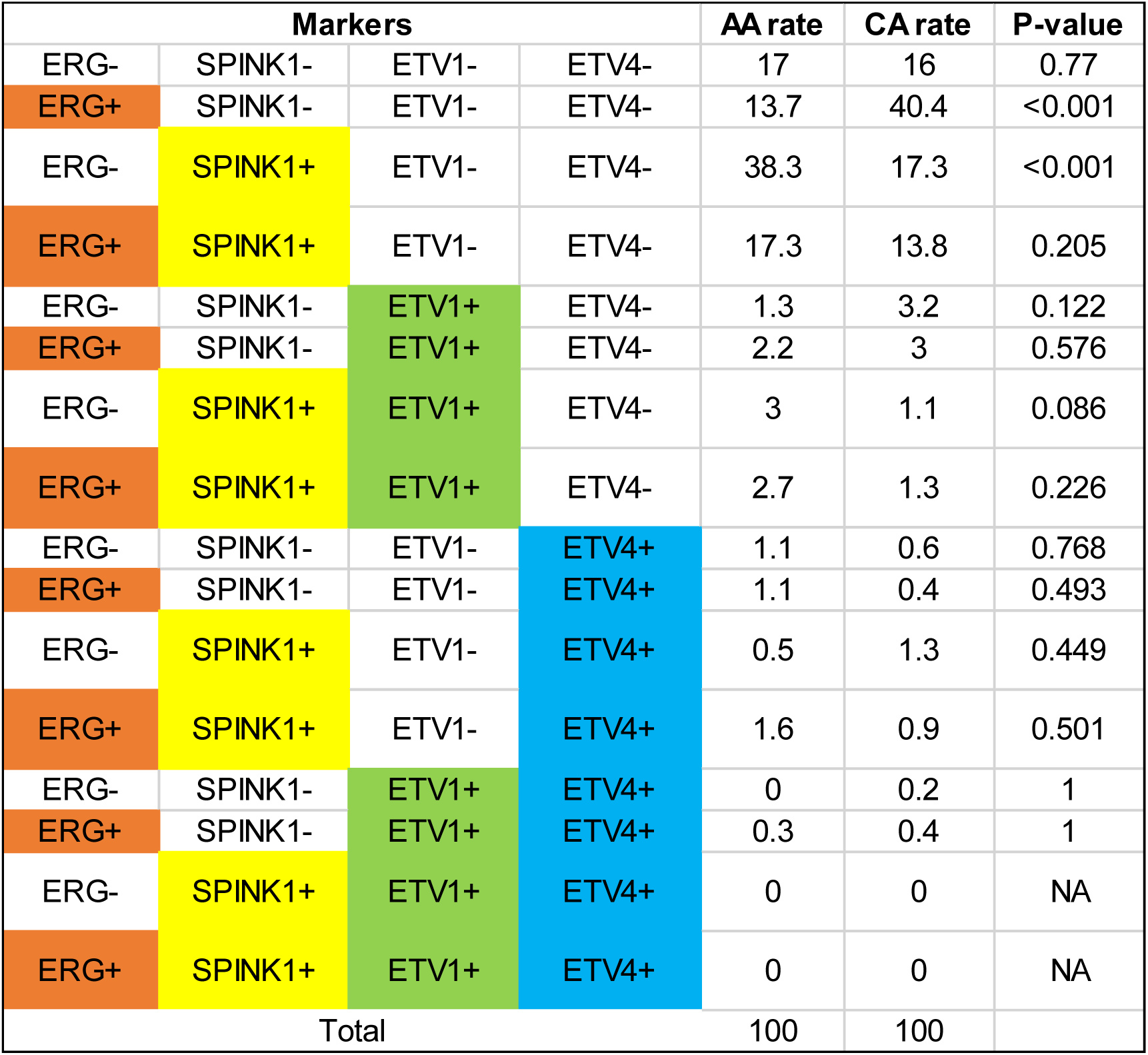
Identification of distinct molecular subtypes of prostate cancer between African Americans (AA) and Caucasian Americans (CA). +: positive expression; -: no expression.

### 3.4 Correlation of oncogene expression with BCR-free survival in AA and CA with localized prostate cancer

In our cohort of 834 patients, 809 individuals underwent routine cancer surveillance after radical prostatectomy, with a median follow-up duration of 4 years. No significant difference in BCR-free survival was observed between AA and CA (log-rank test p=.28), and the median BCR-free survival was not reached (NR). Among the four oncogenes, only ETV4 expression was associated with worse BCR-free survival in AA (log-rank p=.003) (**Figure 4A, S1A-C**). We established a multivariable Cox-PH model to comprehensively evaluate the impact of various prognostic factors. After adjusting for other prognostic factors, ETV4 expression remained significantly associated with worse BCR-free survival in AA (HR = 2.65, 95% CI 1.15-6.09) (**Figure 4B**). Other factors associated with worse BCR-free survival in AA included high pre-operative PSA levels, pathological T3b stage, advanced Gleason Grade groups, regional lymph node metastasis, positive surgical margins, and laterally and posteriorly/posterolaterally located tumors. For CA, only ETV1 expression was marginally associated with worse BCR-free survival (log-rank p=.09) (**Figure 4C, S1D-F**). After adjusting for various prognostic factors, ETV1 expression was significantly associated with worse BCR-free survival in CA (HR=2.36, 95% CI 1.22-4.57) (**Figure 4D**). Other factors associated with worse BCR-free survival in CA included pathological T3b stage, advanced Gleason Grade groups, and positive surgical margins.

**Figure 4:**
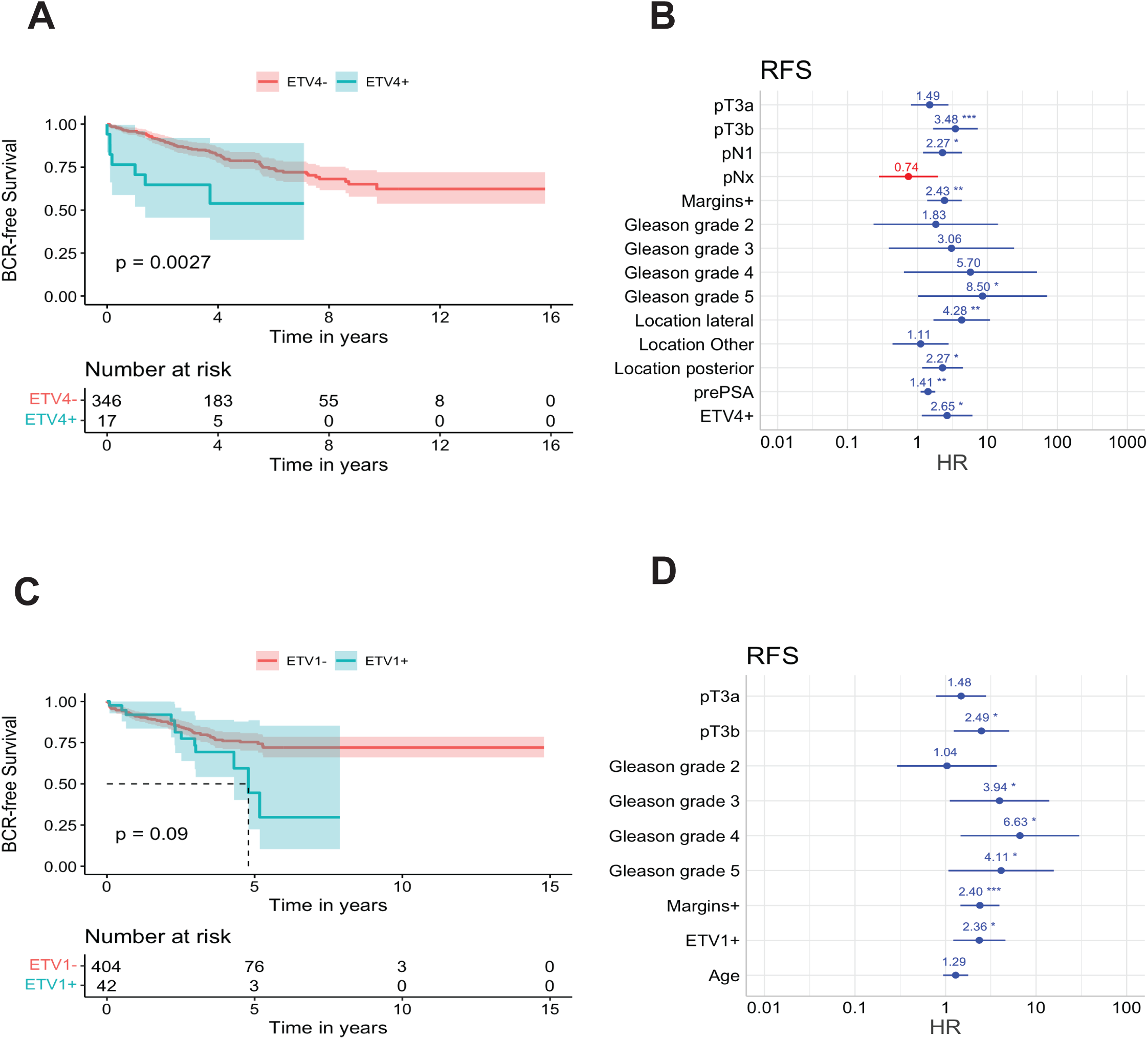
Correlation of Oncogene Expression with Recurrence-free Survival in African American and Caucasian American Patients Following Radical Prostatectomy. (A) Kaplan-Meier curves comparing the recurrence-free survival of African American patients with ETV4 expression (blue line) and those without (red line). (B) Cox-PH models evaluating clinicopathological factors’ hazard ratios and oncogene expression profiles for recurrence-free survival in African American patients. (C) Kaplan-Meier curves comparing the recurrence-free survival between Caucasian American patients with ETV1 expression (blue line) and those without (red line). (D) Cox-PH models evaluating clinicopathological factors’ hazard ratios and oncogene expression profiles for recurrence-free survival in Caucasian American patients. Abbreviations: AA, African American; CA, Caucasian American; HR, hazard ratio; RFS, recurrence-free survival.

### 3.5 Correlation of oncogene expression with regional lymph node metastasis in AA and CA with localized prostate cancer

Among the 708 patients who had pelvic lymph nodes removed during radical prostatectomy, 64 individuals (9.0%) were pathologically confirmed to have lymph node metastasis (pN1). Patients with lymph node metastasis exhibited higher preoperative PSA levels, larger tumor volumes, higher Gleason Grade groups, more advanced pT stages, and more lymphovascular invasion (**Table S5**). Interestingly, the expression of ETV4 was found to be more common in AA with lymph node metastasis compared with those without (OR=5.14%, 95% CI 1.3-17.4, p=.01) (**Figure 5 A, B**). This suggests that ETV4 expression was associated with lymph node metastasis in the context of prostate cancer, particularly in the AA population.

**Figure 5:**
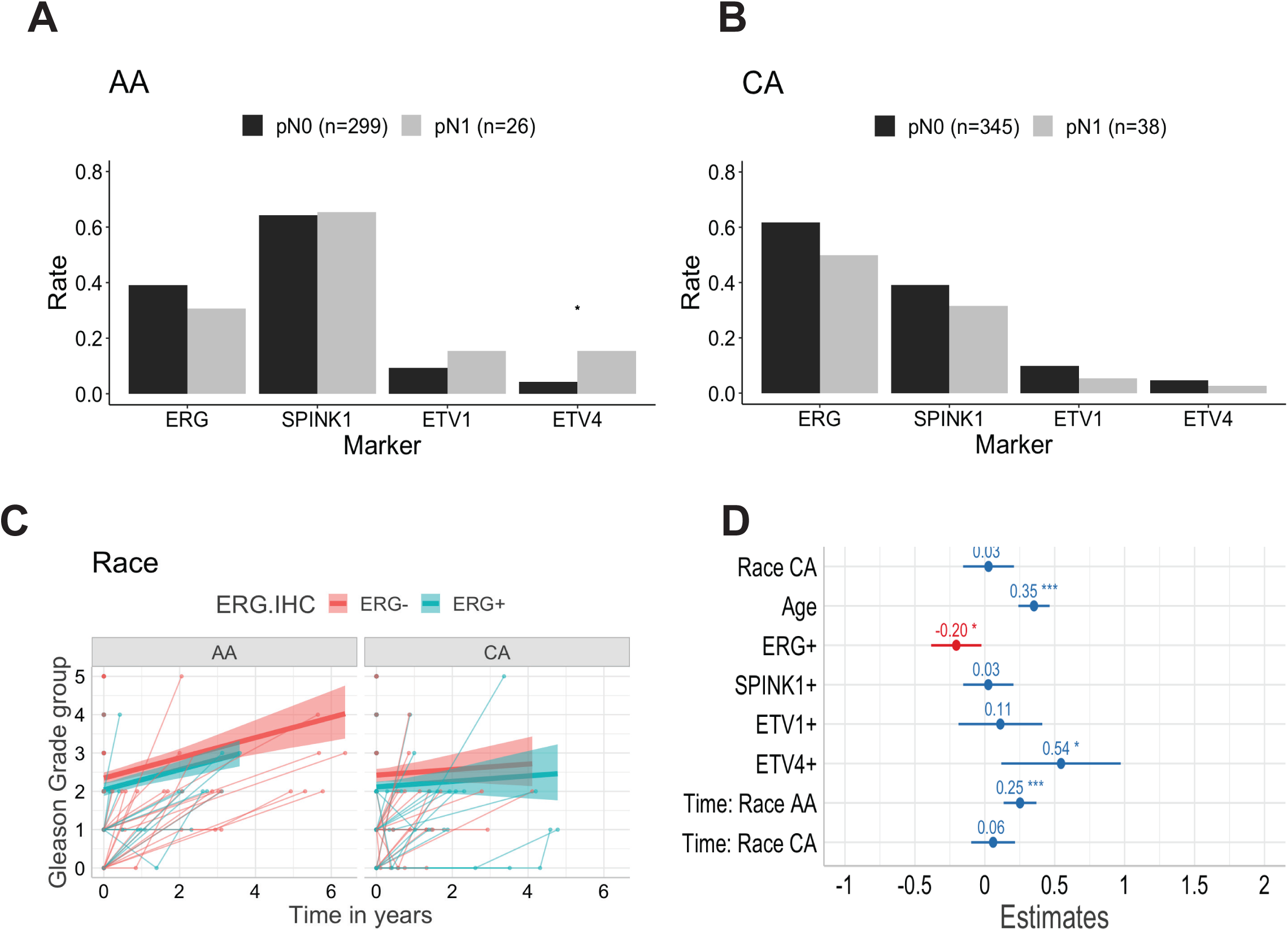
Correlation of Oncogene Expression with Regional Lymph Node Metastasis in African American and Caucasian American Patients at the Time of Radical Prostatectomy. (A) Bar plot comparing expression rates of ERG, SPINK1, ETV1, and ETV4 between African Americans with regional lymph node metastasis and those without at the time of radical prostatectomy. (B) Bar plot comparing expression rates of ERG, SPINK1, ETV1, and ETV4 between Caucasian Americans with regional lymph node metastasis and those without at the time of radical prostatectomy. * p <.05. Temporal Changes in Gleason Grade Group Prior to Radical Prostatectomy. (C) Spaghetti plot summarizing the temporal changes in Gleason Grade groups before radical prostatectomy in African Americans and Caucasian Americans, with thin lines representing individual patients and thick lines representing the mean. Red: patients without ERG expression, blue: patients with ERG expression. (D) Multivariable models evaluate clinicopathological factors’ association and oncogene expression withthe Gleason Grade groups. Abbreviations: AA, African American; CA, Caucasian American

### 3.6 Temporal changes in Gleason Grade group before radical prostatectomy

In our cohort of 834 cases, 726 had one prostate biopsy before prostatectomy, 72 patients had two biopsies, and 17 had three. Notably, the Gleason Grade group showed an increase over time in AA (0.25 per year, p<.001), while no significant change in the Gleason Grade group with time was observed for CA (**Figure 5C**). Moreover, the Gleason Grade group was found to be higher in patients at advanced age (0.35 per 10-year increase, p<.001) and those who expressed ETV4 (0.54, p=.01).

Conversely, patients who expressed ERG showed a lower Gleason Grade group (−0.20, p=.03) (**Figure 5D**). These findings suggest that age and the expression of specific genes, such as ETV4 and ERG, may play a role in determining the Gleason Grade group and potentially the aggressiveness of prostate cancer in these patients.

## 4. DISCUSSION

Prostate cancer in AA is widely recognized for its aggressive nature and association with worse clinical outcomes compared to other racial groups. Several factors contribute to this disparity, including socioeconomic challenges, differences in tumor grade, genomic variations, and tumor location^15^.

Socioeconomic factors such as limited access to healthcare, lower income, and reduced educational opportunities can hinder timely diagnosis and effective treatment^19^. Studies have shown that AA often present with higher Gleason scores, indicating more aggressive tumors at the time of diagnosis^20, 21, 22^. Crucially, research indicates that ensuring equitable access to standardized treatment protocols can significantly improve outcomes for AA patients^23^. For instance, studies have demonstrated that when AA receive similar treatment to their counterparts, including therapies like radical prostatectomy, radiation therapy, and androgen deprivation therapy, their cancer-specific mortality rates are markedly reduced. These findings underscore the importance of addressing both biological and systemic factors to mitigate disparities in prostate cancer outcomes among AA^14^. Consistent with this notion, our study comparing the BCR-free survival in 1207 CA and 994 AA who underwent radical prostatectomy for localized prostate cancer revealed that AA had significantly better outcomes than CA even after adjusting for known nonbiological prognostic factors.

Genomic studies have revealed distinct molecular profiles in prostate tumors from AA men, including a higher prevalence of certain genetic mutations and alterations, such as those involving the ETS gene fusions, SPINK1, and PTEN loss, which are associated with aggressive disease^24, 25^.

Additionally, there are differences in tumor location within the prostate, with AA more frequently having tumors in the anterior prostate, which are more challenging to detect through traditional screening methods^26, 27^. To further understand the involvement of any biological factors that may contribute to the differential clinical outcome, we conducted molecular profiling of a large cohort of wholemount radical prostatectomy cases, including the largest AA cohort. We evaluated three ETS gene fusions and SPINK1 simultaneously on wholemount specimens for the first time using the dual IHC and RNA-ISH approach^2^. The primary objective was to explore the expression patterns of these markers in multifocal prostate cancer and their association with clinical outcomes between AA and CA patients. Conventional approaches revealed controversial observations on the association of these markers with clinical outcomes. Despite the discovery of ETS gene fusions in prostate cancer in 2005^1^, the association of ETS gene fusions with clinical outcomes in prostate cancer has been a topic of debate and controversy. While some studies suggest that ETS gene fusions, such as TMPRSS2-ERG, are associated with more aggressive disease and poorer outcomes, others report conflicting findings^28, 29, 30, 31^. These discrepancies may arise from various factors, including differences in patient cohorts, study methodologies, sampling biases using tissue microarray, analysis limited to index tumor foci, and the specific endpoints analyzed. Notably, no systematic studies have unbiasedly evaluated ETS gene fusions and SPINK1 expression patterns using whole-mount specimens to understand the genetic underpinnings within each tumor focus. In addition to extensive morphological heterogeneity, studies have confirmed significant molecular heterogeneity, with multiple ETS gene fusions and SPINK1 expression present within a single prostate^2, 3, 8, 17^. Although these markers coexist within the prostate, the influence of their combined presence on clinical outcomes remains poorly understood. Our study aims to address this critical clinical question by systematically investigating the impact of ETS gene fusions and SPINK1 expression patterns on prostate cancer outcomes using comprehensive whole-mount specimen analysis.

To better understand the role of ETS gene fusions in prostate cancer outcomes, we conducted an integrated approach combining molecular analyses with clinical data. We identified subgroups of patients with more than one ETS gene fusion within a prostate along with SPINK1. The presence of more than one driver molecular aberration is a rare and unique phenomenon not reported previously. Therefore, this study marks the first attempt to characterize molecular subtypes in these racial groups, considering both dominant and more minor foci of prostate cancer. Our research findings reaffirmed the multifocality of prostate cancer, indicating that each tumor focus may exhibit distinct molecular aberrations and histological patterns. Notably, we identified cases with tumors expressing single-driver mutations as well as cases with two or three-driver mutations that were mutually exclusive within individual prostate foci but coexisted across different foci in the same specimen. We previously reported a study utilizing dual IHC and dual RNA-ISH to evaluate 601 biopsy cores (standard 12-core biopsy) from 120 patients. This study revealed that 45% of the cores were positive for at least one of the molecular markers (ERG, SPINK1, ETV1, ETV4), with ERG being the most prominent marker. Notably, cases of dual marker expression were also observed, but these were exclusive to separate tumor foci, suggesting different clonal origins^2^. This study further highlights the molecular heterogeneity and the mutual exclusivity of these markers within single tumor foci, aligning with our observations in whole-mount radical prostatectomy specimens. Another study focused on the clonal evaluation of early-onset prostate cancer through molecular profiling of these markers on whole-mount radical prostatectomy tissue, corroborating the clonal heterogeneity of prostate cancer^3^. These studies further highlight the molecular heterogeneity and the mutual exclusivity of these markers within single tumor foci, aligning with our current observations in whole-mount radical prostatectomy (RP) specimens. Even though we have identified new molecular subsets, approximately 17% of the cases are negative for all four markers evaluated. This is mainly because we selected only one of the many tissue blocks containing cancer for each patient. Each patient typically has at least 5-7 tissue blocks available. Future studies that evaluate all tumor blocks with cancer may eliminate or the numbers reduced in the subset of cases that are negative for the ETS gene fusion and SPINK1. In a related study using saturation biopsies (>20 biopsy cores obtained for each patient) we observed about 99% of the cases were positive for at least one of the ETS gene fusions or SPINK1 and large subset positive with two-four markers. (Palanisamy et al., unpublished data).

These findings contribute to the understanding of tumor heterogeneity and the specific roles of these oncogenes in prostate cancer. Moreover, the expression and implications of ERG in prostate cancer have been extensively studied, with its role in diagnosis and potential prognostic value being noted. The complex relationship between ERG expression and prostate cancer progression is a significant area of research, offering insights into molecular subtyping and potential therapeutic targets. These studies collectively contribute to a deeper understanding of prostate cancer’s molecular complexity and heterogeneity, especially concerning the expressions of ERG, ETV1, ETV4, and SPINK1. They provide a substantial basis for further research and potential clinical applications, particularly in clonal evaluation and targeted therapy strategies. Integrating these findings, the results of this present study emphasize the significance of studying the mutual exclusivity and coexistence of these oncogenes in prostate cancer, offering a comprehensive view of their roles and implications.

Future studies with additional markers including ETV5, SPOP, FOXA1 and others using multiplex screening approaches may reveal the molecular underpinnings in tumor foci that are negative for ERG, ETV1, ETV4 and SPINK1.

This novel insight, often overlooked in previous research, was made possible by our unique sampling approach. Additionally, we investigated the connection between the molecular profile of radical prostatectomy specimens and patients’ likelihood of BCR. Our objective was to understand whether racial variations in tumor genomics contribute to the observed racial disparity in prostate cancer outcomes. In line with previous studies, we found that the expression of ERG and SPINK1 had limited prognostic value in predicting cancer recurrence following radical prostatectomy. However, our investigation revealed significant associations between ETV1 expression and a higher BCR incidence in CA patients and between ETV4 expression and a higher BCR incidence in AA patients, even after accounting for other prognostic factors. These findings suggest that ETV1 and ETV4 expression might be potential markers for predicting BCR risk in CA and AA patients after radical prostatectomy, respectively. Overall, this study sheds light on the complex molecular heterogeneity of prostate cancer in different racial groups and offers valuable insights into potential prognostic factors for BCR in these patients.

For the first time, we showed that ETV1 and ETV4 expression are associated with worse BCR-free survival, highlighting a racial disparity. This finding is significant because, until now, no studies have conducted molecular profiling for ETV1 and ETV4 using whole-mount specimens in conjunction with ERG and SPINK1. One reason for this gap is the lack of cancer-specific antibodies for ETV1 and ETV4. Our study used RNA-ISH for ETV1 and ETV4 and IHC for ERG and SPINK1. We have reported the concordance of ETV1 and ETV4 positivity by RNA-ISH with fluorescence in situ hybridization (FISH) to confirm rearrangement in these genes^32^ as these genes are expressed only with the result of chromosome translocation. Previous studies primarily used fluorescence in situ hybridization (FISH) for ETV1 and ETV4 separately, and these analyses were mostly performed on tissue microarrays (TMA) or selected tumor foci from radical prostatectomy specimens. Our approach provides more comprehensive and unbiased molecular profiling by preserving the spatial distribution of tumors, which may explain the newly observed racial disparities in BCR-free survival associated with ETV1 and ETV4 expression.

Our study has provided compelling evidence indicating that identical genetic aberrations in men diagnosed with localized prostate cancer can have significantly different prognostic values depending on their race. One plausible explanation for this observation is the existence of racial differences in the cancer genome among prostate cancer patients, which can influence the biological impact of specific genetic aberrations. It is well recognized that the ectopic expression of ETV1 or ETV4, driven by gene rearrangement, is insufficient to initiate tumorigenesis in prostate tissue. Instead, these gene rearrangements require synergistic interactions with other concurrent genetic aberrations, such as PTEN loss and TP53 mutations. It has been shown that the introduction of N terminus-truncated human ERG or ETV1 into the TMPRSS2 locus did not lead to the development of prostate adenocarcinoma in genetically engineered mouse (GEM) models with heterogeneous loss of PTEN. However, when the TMPRSS2-ETV1 fusion was introduced into GEM models with homogenous loss of PTEN, it resulted in the rapid development of invasive prostate adenocarcinoma^4^. It has also been shown that the interplay between ETV1 and PTEN enhanced the oncogenic capacity of ETV1 when there was concurrent partial loss of PTEN^33^. Therefore, the differential prognostic values of ETV1 in CA and AA with localized prostate cancer may be attributed, in part, to a higher incidence of PTEN loss in CA compared to AA, as reported in several studies. On the other hand, research by Li et al.^7^ revealed that ETV4 gene rearrangement significantly upregulated TP53 and genes associated with p53-induced senescence. Consequently, the oncogenic transformation of normal prostate epithelium following ETV4 gene rearrangement was hindered by simultaneous senescence activation, leading to the death of mutated cells that would have otherwise become invasive adenocarcinoma. In a GEM model, ETV4 overexpression, in cooperation with TP53 loss, induced diffuse neoplasia of prostate tissue, while ETV4 overexpression alone led to prostatic intraepithelial neoplasia that later regressed over time. It would be reasonable to assume that ETV4 expression in prostate cancer would confer more prognostic significance in CA, given a higher prevalence of TP53 mutations observed in this racial group. Contrary to this assumption, our study showed that ETV4 expression was associated with BCR exclusively in AA. Since prostate cancer in AA has a higher prevalence of MYC amplification compared to CA, this discrepancy might be explained by the suppression of ETV4-induced senescence by MYC amplification in a p53-independent mechanism^34, 35^. Therefore, it is essential to recognize that these findings represent only the beginning of our journey to understanding how prostate cancer initiates, progresses, and relapses. Until the distant future, when all genetic aberrations associated with prostate cancer and their complex interactions are fully uncovered, race will continue to serve as a practical surrogate to account for the unique genomic background of each individual.

Remarkably, current commercially available molecular tests for prostate cancer have not yet integrated race as a factor in their algorithms to customize molecular panels for individual patients. One such test is the Decipher Genomic Classifier, which relies on the expression of 22 genes, including coding and noncoding RNA markers and does not evaluate the presence of any of the ETS gene fusions and SPINK1 in high Gleason-grade tumors or consider both dominant and secondary tumor foci^36^. This test categorizes patients with localized prostate cancer into low, intermediate, or high risk of early metastasis. It has been shown to correlate with prostate cancer-specific mortality and overall survival after metastasis. Developing the Decipher Genomic Classifier involved comparing the RNA profiles of patients who developed early regional or distant metastasis confirmed by imaging with those with no recurrence or BCR. However, the authors did not consider race, and the race composition of their study cohort was not reported. Consequently, despite offering personalized prognostication, the utility of the Decipher Genomic Classifier in guiding clinical decisions following radical prostatectomy still needs to be more conclusive. To further enhance its clinical relevance and accuracy, future research should evaluate the performance of the Decipher Genomic Classifier, specifically in AA and CA populations. By doing so, we can determine if there are racial disparities in its predictive capabilities. Moreover, assessing whether incorporating ETV1 and ETV4 markers, as discussed in our study, would provide additional prognostic value in these populations would be valuable. This approach could help tailor treatment plans and improve outcomes for prostate cancer patients of different racial backgrounds.

In conclusion, our study revealed that AA patients diagnosed with localized prostate cancer and expressing ETV4 experienced a higher incidence of BCR following radical prostatectomy. Conversely, CA patients diagnosed with localized prostate cancer and expressing ETV1 had a higher incidence of BCR following radical prostatectomy. These findings underscore the critical importance of considering racial differences in interpreting molecular data and developing genomic profiles, ensuring precise and personalized prognostication. Using a single wholemount tissue block in our research has provided significant insights into the molecular underpinnings of prostate cancer. Future studies evaluating every wholemount tissue block with cancer from each patient will significantly reduce or eliminate negative cases for ETS gene fusions and SPINK1. This approach will uncover distinct molecular subsets of cases associated with clinical outcomes. Our unpublished data using saturation biopsy cases revealed that about 99% of the cases are positive for one or more of the ETS gene fusion and SPINK1, suggesting the rationale for saturation biopsy screening rather than the standard 12-core biopsy approach. This approach has enabled a more comprehensive analysis compared to standard twelve-core biopsies, highlighting the potential for more accurate identification of molecular subtypes with prognostic significance. Consequently, future studies should focus on molecular profiling using saturation biopsies rather than standard biopsies to capture the heterogeneity of the tumor more effectively rather than conveniently missing valuable prognostic information at diagnosis. Such advancements in molecular profiling at the time of diagnosis would aid clinicians and patients in making informed decisions regarding treatment options and follow-up strategies. By identifying distinct molecular subtypes with prognostic markers, personalized treatment plans can be developed, potentially improving clinical outcomes. This approach aligns with the growing emphasis on precision medicine, where treatment is tailored to the individual characteristics of each patient’s disease, ultimately leading to better management and prognosis of prostate cancer.

## Data Availability

we will make the raw data and images available

## Acknowledgments

We thank Jingli Yang, Natalia Draga, Jessica Ryba, and Hristina Trpevski for their help in preparing slides from whole-mount tissue blocks. This study was supported by a US Department of Defense grant W81XWH-16-1-0544 to Nallasivam Palanisamy and internal funding from the Henry Ford Health System research administration.

