## supplement tables and figures for ""Unveiling Prostate Cancer’s Molecular Tapestry: Ethnic Disparities and Prognostic Insights from Whole-Mount Prostatectomy Tissue Analysis""

**Table S1.** Comparison of Patients with Localized Prostate Cancer by ERG Expression Status

| | | ERG- | ERG+ | Total | p* |
|---|---|---|---|---|---|
| **Total N (%)** | | 411 (49.3) | 423 (50.7) | 834 | |
| **Race** | AA | 227 (55.2) | 144 (34.0) | 371 (44.5) | <0.001 |
| | CA | 184 (44.8) | 279 (66.0) | 463 (55.5) | |
| **Age (years)** | Median (IQR) | 63.0 (57.0 to 68.0) | 60.0 (55.0 to 65.0) | 61.0 (56.0 to 67.0) | <0.001 |
| **Preoperative PSA (ng/mL)** | Median (IQR) | 5.8 (4.5 to 8.7) | 5.4 (4.4 to 7.3) | 5.6 (4.4 to 8.1) | 0.023 |
| **Family history** | Negative | 134 (32.6) | 134 (31.7) | 268 (32.1) | 0.228 |
| | Other cancer | 128 (31.1) | 112 (26.5) | 240 (28.8) | |
| | Prostate cancer | 126 (30.7) | 142 (33.6) | 268 (32.1) | |
| | Unknown | 23 (5.6) | 35 (8.3) | 58 (7.0) | |
| **Tumor volume (%)** | Median (IQR) | 10.0 (5.0 to 18.0) | 10.0 (5.0 to 15.0) | 10.0 (5.0 to 7.0) | 0.683 |
| **Tumor stage** | pT2 | 225 (55.0) | 241 (57.1) | 466 (56.1) | 0.31 |
| | pT3a | 125 (30.6) | 135 (32.0) | 260 (31.3) | |
| | pT3b | 59 (14.4) | 46 (10.9) | 105 (12.6) | |
| **Node stage** | pN0 | 314 (76.4) | 330 (78.0) | 644 (77.2) | 0.355 |
| | pN1 | 37 (9.0) | 27 (6.4) | 64 (7.7) | |
| | pNx | 60 (14.6) | 66 (15.6) | 126 (15.1) | |
| **Gleason Grade group** | 1 | 47 (11.6) | 55 (13.0) | 102 (12.3) | 0.017 |
| | 2 | 191 (47.3) | 235 (55.7) | 426 (51.6) | |
| | 3 | 104 (25.7) | 96 (22.7) | 200 (24.2) | |
| | 4 | 17 (4.2) | 11 (2.6) | 28 (3.4) | |
| | 5 | 45 (11.1) | 25 (5.9) | 70 (8.5) | |
| **Tumor margins** | Negative | 270 (65.7) | 283 (66.9) | 553 (66.3) | 0.767 |
| | Positive | 141 (34.3) | 140 (33.1) | 281 (33.7) | |
| **Perineural invasion** | Absent | 237 (57.7) | 239 (56.5) | 476 (57.1) | 0.788 |

| | | | | | |
|---|---|---|---|---|---|
| | Present | 174 (42.3) | 184 (43.5) | 358 (42.9) | |
| **Lymph vascular invasion** | Absent | 357 (86.9) | 378 (89.4) | 735 (88.1) | 0.313 |
| | Present | 54 (13.1) | 45 (10.6) | 99 (11.9) | |
| **Tumor location** | Anterior/anterolateral | 121 (29.4) | 72 (17.0) | 193 (23.1) | <0.001 |
| | Lateral | 36 (8.8) | 46 (10.9) | 82 (9.8) | |
| | Other | 57 (13.9) | 51 (12.1) | 108 (12.9) | |
| | Posterior/posterolateral | 197 (47.9) | 254 (60.0) | 451 (54.1) | |

* Chi-squared test for categorical variables, Wilcoxon rank test for continuous variables. Abbreviations: AA, African American; CA, Caucasian American; IQR, interquartile range; PSA, prostate-specific antigen.

**Table S2.** Comparison of Patients with Localized Prostate Cancer by SPINK1 Expression Status

| | | SPINK1- | SPINK1+ | Total | p* |
|---|---|---|---|---|---|
| **Total N (%)** | | 434 (52.0) | 400 (48.0) | 834 | |
| **Race** | AA | 136 (31.3) | 235 (58.8) | 371 (44.5) | <0.001 |
| | CA | 298 (68.7) | 165 (41.2) | 463 (55.5) | |
| **Age (years)** | Median (IQR) | 61.0 (56.0 to 67.0) | 61.0 (56.0 to 7.0) | 61.0 (56.0 to 7.0) | 0.88 |
| **Preoperative PSA (ng/mL)** | Median (IQR) | 5.3 (4.2 to 7.6) | 5.9 (4.6 to 8.5) | 5.6 (4.4 to 8.1) | 0.001 |
| **Family history** | Negative | 130 (30.0) | 138 (34.5) | 268 (32.1) | 0.339 |
| | Other cancer | 134 (30.9) | 106 (26.5) | 240 (28.8) | |
| | Prostate cancer | 137 (31.6) | 131 (32.8) | 268 (32.1) | |
| | Unknown | 33 (7.6) | 25 (6.2) | 58 (7.0) | |
| **Tumor volume (%)** | Median (IQR) | 8.0 (4.0 to 15.0) | 10.0 (6.0 to 20.0) | 10.0 (5.0 to 17.0) | <0.001 |
| **Tumor stage** | pT2 | 257 (59.5) | 209 (52.4) | 466 (56.1) | 0.067 |
| | pT3a | 120 (27.8) | 140 (35.1) | 260 (31.3) | |
| | pT3b | 55 (12.7) | 50 (12.5) | 105 (12.6) | |
| **Node stage** | pN0 | 317 (73.0) | 327 (81.8) | 644 (77.2) | 0.004 |
| | pN1 | 35 (8.1) | 29 (7.2) | 64 (7.7) | |
| | pNx | 82 (18.9) | 44 (11.0) | 126 (15.1) | |
| **Gleason Grade Group** | 1 | 70 (16.4) | 32 (8.0) | 102 (12.3) | <0.001 |
| | 2 | 215 (50.4) | 211 (52.9) | 426 (51.6) | |
| | 3 | 85 (19.9) | 115 (28.8) | 200 (24.2) | |
| | 4 | 11 (2.6) | 17 (4.3) | 28 (3.4) | |
| | 5 | 46 (10.8) | 24 (6.0) | 70 (8.5) | |
| **Tumor margins** | Negative | 298 (68.7) | 255 (63.8) | 553 (66.3) | 0.154 |
| | Positive | 136 (31.3) | 145 (36.2) | 281 (33.7) | |
| **Perineural invasion** | Absent | 259 (59.7) | 217 (54.2) | 476 (57.1) | 0.131 |
| | Present | 175 (40.3) | 183 (45.8) | 358 (42.9) | |
| **Lymph vascular invasion** | Absent | 378 (87.1) | 357 (89.2) | 735 (88.1) | 0.393 |
| | Present | 56 (12.9) | 43 (10.8) | 99 (11.9) | |
| **Tumor location** | Anterior/anterolateral | 82 (18.9) | 111 (27.8) | 193 (23.1) | 0.02 |
| | Lateral | 44 (10.1) | 38 (9.5) | 82 (9.8) | |
| | Other | 63 (14.5) | 45 (11.2) | 108 (12.9) | |
| | Posterior/posterolateral | 245 (56.5) | 206 (51.5) | 451 (54.1) | |

* Chi-squared test for categorical variables, Wilcoxon rank test for continuous variables. Abbreviations: AA, African American; CA, Caucasian American; IQR, interquartile range; PSA, prostate-specific antigen.

**Table S3.** Comparison of Patients with Localized Prostate Cancer by ETV1 Expression Status

| | | ETV1- | ETV1+ | Total | p* |
|---|---|---|---|---|---|
| **Total N (%)** | | 756 (90.6) | 78 (9.4) | 834 | |
| **Race** | AA | 336 (44.4) | 35 (44.9) | 371 (44.5) | 1 |
| | CA | 420 (55.6) | 43 (55.1) | 463 (55.5) | |
| **Age (years)** | Median (IQR) | 62.0 (56.0 to 67.0) | 60.5 (57.2 to 64.0) | 61.0 (56.0 to 67.0) | 0.531 |
| **Preoperative PSA (ng/mL)** | Median (IQR) | 5.5 (4.4 to 7.9) | 5.8 (4.5 to 9.5) | 5.6 (4.4 to 8.1) | 0.269 |
| **Family history** | Negative | 243 (32.1) | 25 (32.1) | 268 (32.1) | 0.238 |
| | Other cancer | 220 (29.1) | 20 (25.6) | 240 (28.8) | |
| | Prostate cancer | 237 (31.3) | 31 (39.7) | 268 (32.1) | |
| | Unknown | 56 (7.4) | 2 (2.6) | 58 (7.0) | |
| **Tumor volume (%)** | Median (IQR) | 10.0 (5.0 to 16.0) | 10.0 (7.0 to 25.0) | 10.0 (5.0 to 17.0) | 0.013 |
| **Tumor stage** | pT2 | 431 (57.2) | 35 (44.9) | 466 (56.1) | 0.065 |
| | pT3a | 232 (30.8) | 28 (35.9) | 260 (31.3) | |
| | pT3b | 90 (12.0) | 15 (19.2) | 105 (12.6) | |
| **Node stage** | pN0 | 582 (77.0) | 62 (79.5) | 644 (77.2) | 0.837 |
| | pN1 | 58 (7.7) | 6 (7.7) | 64 (7.7) | |
| | pNx | 116 (15.3) | 10 (12.8) | 126 (15.1) | |
| **Gleason Grade group** | 1 | 92 (12.3) | 10 (12.8) | 102 (12.3) | 0.451 |
| | 2 | 383 (51.2) | 43 (55.1) | 426 (51.6) | |
| | 3 | 183 (24.5) | 17 (21.8) | 200 (24.2) | |
| | 4 | 28 (3.7) | 0 (0.0) | 28 (3.4) | |
| | 5 | 62 (8.3) | 8 (10.3) | 70 (8.5) | |
| **Tumor margins** | Negative | 504 (66.7) | 49 (62.8) | 553 (66.3) | 0.577 |
| | Positive | 252 (33.3) | 29 (37.2) | 281 (33.7) | |
| **Perineural invasion** | Absent | 426 (56.3) | 50 (64.1) | 476 (57.1) | 0.231 |
| | Present | 330 (43.7) | 28 (35.9) | 358 (42.9) | |
| **Lymph vascular invasion** | Absent | 668 (88.4) | 67 (85.9) | 735 (88.1) | 0.648 |
| | Present | 88 (11.6) | 11 (14.1) | 99 (11.9) | |
| **Tumor location** | Anterior/anterolateral | 175 (23.1) | 18 (23.1) | 193 (23.1) | 0.917 |
| | Lateral | 76 (10.1) | 6 (7.7) | 82 (9.8) | |
| | Other | 97 (12.8) | 11 (14.1) | 108 (12.9) | |
| | Posterior/posterolateral | 408 (54.0) | 43 (55.1) | 451 (54.1) | |

* Chi-squared test for categorical variables, Wilcoxon rank test for continuous variables. Abbreviations: AA, African American; CA, Caucasian American; IQR, interquartile range; PSA, prostate-specific antigen.

**Table S4.** Comparison of Patients with Localized Prostate Cancer by *ETV4* Expression Status

| | | ETV4- | ETV4+ | Total | p* |
|---|---|---|---|---|---|
| **Total N (%)** | | 799 (95.8) | 35 (4.2) | 834 | |
| **Race** | AA | 354 (44.3) | 17 (48.6) | 371 (44.5) | 0.746 |
| | CA | 445 (55.7) | 18 (51.4) | 463 (55.5) | |
| **Age (years)** | Median (IQR) | 61.0 (56.0 to 67.0) | 60.0 (55.0 to 70.0) | 61.0 (56.0 to 67.0) | 0.769 |
| **Preoperative PSA (ng/mL)** | Median (IQR) | 5.5 (4.4 to 8.0) | 6.8 (4.9 to 8.9) | 5.6 (4.4 to 8.1) | 0.044 |
| **Family history** | Negative | 258 (32.3) | 10 (28.6) | 268 (32.1) | 0.13 |
| | Other cancer | 224 (28.0) | 16 (45.7) | 240 (28.8) | |
| | Prostate cancer | 260 (32.5) | 8 (22.9) | 268 (32.1) | |
| | Unknown | 57 (7.1) | 1 (2.9) | 58 (7.0) | |
| **Tumor volume (%)** | Median (IQR) | 10.0 (5.0 to 16.0) | 7.0 (4.0 to 20.0) | 10.0 (5.0 to 17.0) | 0.419 |
| **Tumor stage** | pT2 | 445 (55.8) | 21 (61.8) | 466 (56.1) | 0.13 |
| | pT3a | 254 (31.9) | 6 (17.6) | 260 (31.3) | |
| | pT3b | 98 (12.3) | 7 (20.6) | 105 (12.6) | |
| **Node stage** | pN0 | 615 (77.0) | 29 (82.9) | 644 (77.2) | 0.053 |
| | pN1 | 59 (7.4) | 5 (14.3) | 64 (7.7) | |
| | pNx | 125 (15.6) | 1 (2.9) | 126 (15.1) | |
| **Gleason Grade Group** | 1 | 101 (12.8) | 1 (2.9) | 102 (12.3) | 0.153 |
| | 2 | 411 (52.0) | 15 (42.9) | 426 (51.6) | |
| | 3 | 187 (23.6) | 13 (37.1) | 200 (24.2) | |
| | 4 | 26 (3.3) | 2 (5.7) | 28 (3.4) | |
| | 5 | 66 (8.3) | 4 (11.4) | 70 (8.5) | |
| **Tumor margins** | Negative | 535 (67.0) | 18 (51.4) | 553 (66.3) | 0.085 |
| | Positive | 264 (33.0) | 17 (48.6) | 281 (33.7) | |
| **Perineural invasion** | Absent | 450 (56.3) | 26 (74.3) | 476 (57.1) | 0.054 |
| | Present | 349 (43.7) | 9 (25.7) | 358 (42.9) | |
| **Lymph vascular invasion** | Absent | 706 (88.4) | 29 (82.9) | 735 (88.1) | 0.473 |
| | Present | 93 (11.6) | 6 (17.1) | 99 (11.9) | |
| **Tumor location** | Anterior/anterolateral | 187 (23.4) | 6 (17.1) | 193 (23.1) | 0.208 |
| | Lateral | 77 (9.6) | 5 (14.3) | 82 (9.8) | |
| | Other | 100 (12.5) | 8 (22.9) | 108 (12.9) | |
| | Posterior/posterolateral | 435 (54.4) | 16 (45.7) | 451 (54.1) | |

* Chi-squared test for categorical variables, Wilcoxon rank test for continuous variables. Abbreviations: AA, African American; CA, Caucasian American; IQR, interquartile range; PSA, prostate-specific antigen.

**Table S5.** Comparison of Patients with or without Regional Nodal Metastasis

| | | pN0 | pN1 | Total | p* |
|---|---|---|---|---|---|
| **Total N (%)** | | 644 (91.0) | 64 (9.0) | 708 | |
| **Race** | AA | 299 (46.4) | 26 (40.6) | 325 (45.9) | 0.449 |
| | CA | 345 (53.6) | 38 (59.4) | 383 (54.1) | |
| **ERG** | Negative | 314 (48.8) | 37 (57.8) | 351 (49.6) | 0.211 |
| | Positive | 330 (51.2) | 27 (42.2) | 357 (50.4) | |
| **SPINK1** | Negative | 317 (49.2) | 35 (54.7) | 352 (49.7) | 0.482 |
| | Positive | 327 (50.8) | 29 (45.3) | 356 (50.3) | |
| **ETV1** | Negative | 582 (90.4) | 58 (90.6) | 640 (90.4) | 1 |
| | Positive | 62 (9.6) | 6 (9.4) | 68 (9.6) | |
| **ETV4** | Negative | 615 (95.5) | 59 (92.2) | 674 (95.2) | 0.382 |
| | Positive | 29 (4.5) | 5 (7.8) | 34 (4.8) | |
| **Age (years)** | Median (IQR) | 61.0 (56.0 to 67.0) | 63.0 (57.0 to 69.0) | 61.5 (56.0 to 67.0) | 0.067 |
| **Preoperative PSA (ng/mL)** | Median (IQR) | 5.7 (4.5 to 8.2) | 8.2 (5.5 to 14.0) | 5.9 (4.6 to 8.5) | <0.001 |
| **Family history** | Negative | 202 (31.4) | 25 (39.1) | 227 (32.1) | 0.284 |
| | Other cancer | 194 (30.1) | 22 (34.4) | 216 (30.5) | |
| | Prostate cancer | 201 (31.2) | 13 (20.3) | 214 (30.2) | |
| | Unknown | 47 (7.3) | 4 (6.2) | 51 (7.2) | |
| **Tumor volume (%)** | Median (IQR) | 10.0 (5.0 to 16.0) | 22.0 (10.0 to 45.0) | 10.0 (5.0 to 18.0) | <0.001 |
| **Gleason Grade group** | 1 | 55 (8.5) | 0 (0.0) | 55 (7.8) | <0.001 |
| | 2 | 356 (55.3) | 8 (12.5) | 364 (51.4) | |
| | 3 | 162 (25.2) | 23 (35.9) | 185 (26.1) | |
| | 4 | 23 (3.6) | 4 (6.2) | 27 (3.8) | |
| | 5 | 41 (6.4) | 28 (43.8) | 69 (9.7) | |
| **Tumor stage** | pT2 | 362 (56.2) | 3 (4.7) | 365 (51.6) | <0.001 |
| | pT3a | 219 (34.0) | 19 (29.7) | 238 (33.6) | |
| | pT3b | 61 (9.5) | 41 (64.1) | 102 (14.4) | |
| **Tumor margins** | Negative | 436 (67.7) | 19 (29.7) | 455 (64.3) | <0.001 |
| | Positive | 208 (32.3) | 45 (70.3) | 253 (35.7) | |
| **Perineural invasion** | Absent | 377 (58.5) | 41 (64.1) | 418 (59.0) | 0.469 |

| | | | | | |
|---|---|---|---|---|---|
| | Present | 267 (41.5) | 23 (35.9) | 290 (41.0) | |
| **Lymph vascular invasion** | Absent | 599 (93.0) | 14 (21.9) | 613 (86.6) | <0.001 |
| | Present | 45 (7.0) | 50 (78.1) | 95 (13.4) | |
| **Tumor location** | Anterior/anterolateral | 155 (24.1) | 5 (7.8) | 160 (22.6) | 0.001 |
| | Lateral | 65 (10.1) | 5 (7.8) | 70 (9.9) | |
| | Other | 69 (10.7) | 16 (25.0) | 85 (12.0) | |
| | Posterior/posterolateral | 355 (55.1) | 38 (59.4) | 393 (55.5) | |

* Chi-squared test for categorical variables, Wilcoxon rank test for continuous variables. Abbreviations: AA, African American; CA, Caucasian American; IQR, interquartile range; PSA, prostate-specific antigen.

Zhao et al., Figure S1

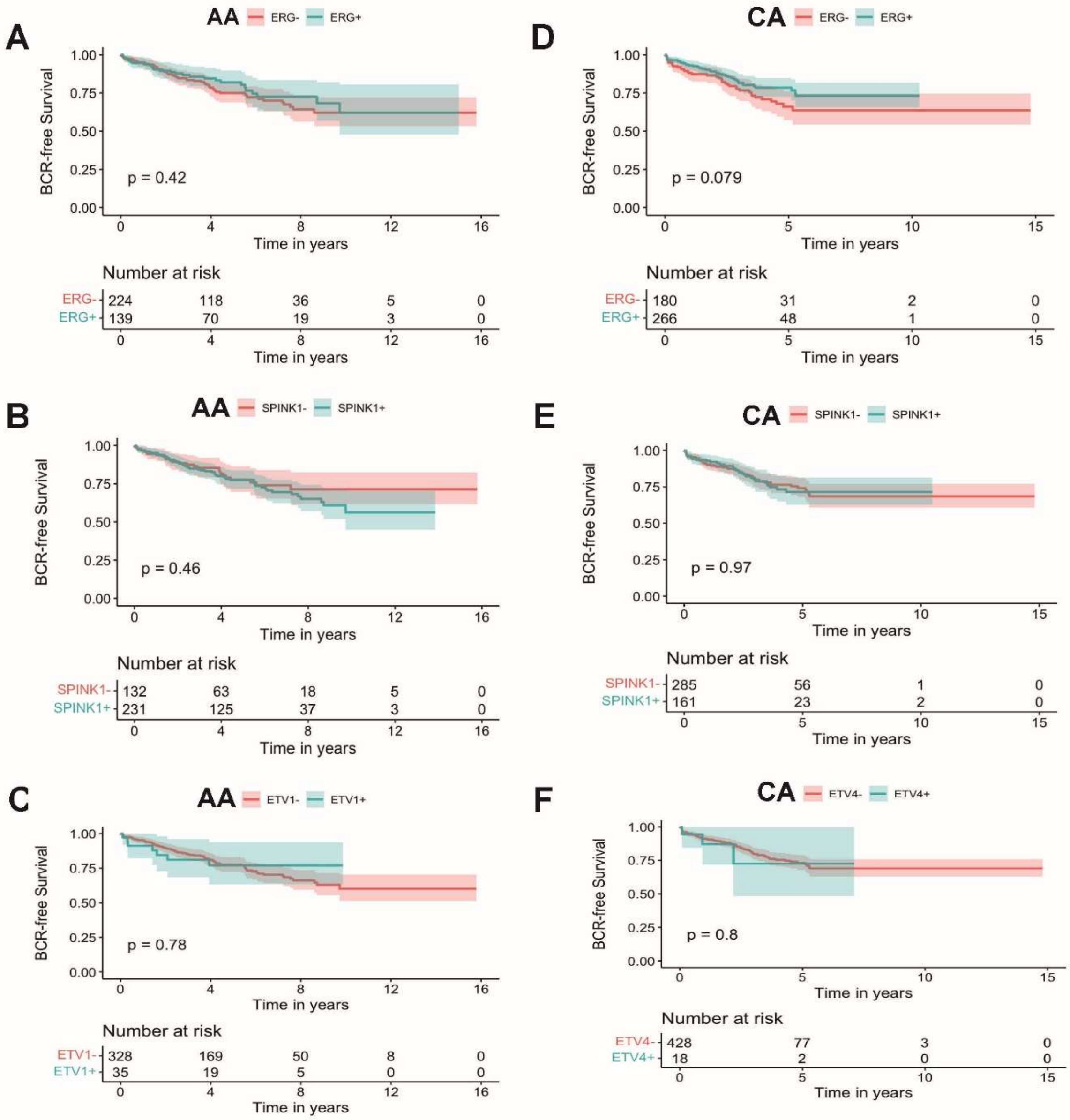


**Figure S1:**Correlation of Oncogene Expression with Recurrence-free Survival in African American( A-C) and Caucasian American (D-F). Patients Following Radical Prostatectomy. (A) Kaplan-Meier curves comparing the recurrence-free survival of African American patients with ERG expression (blue line) and those without (red line). (B) Kaplan-Meier curves comparing the recurrence-free survival of African American patients with SPINK1 expression (blue line) and those without (red line). (C) Kaplan-Meier curves comparing the recurrence-free survival of African American patients with ETV1 expression (blue line) and those without (red line). (D) Kaplan-Meier curves comparing the recurrence-free survival of African American patients with ERG expression (blue line) and those without (red line). (E) Kaplan-Meier curves comparing the recurrence-free survival of African American patients with SPINK1 expression (blue line) and those without (red line). (F) Kaplan-Meier curves comparing the recurrence-free survival of African American patients with ETV4 expression (blue line) and those without (red line). Abbreviations: AA, African American; CA, Caucasian American; RFS, recurrence-free survival.

Zhao et al., Figure S2

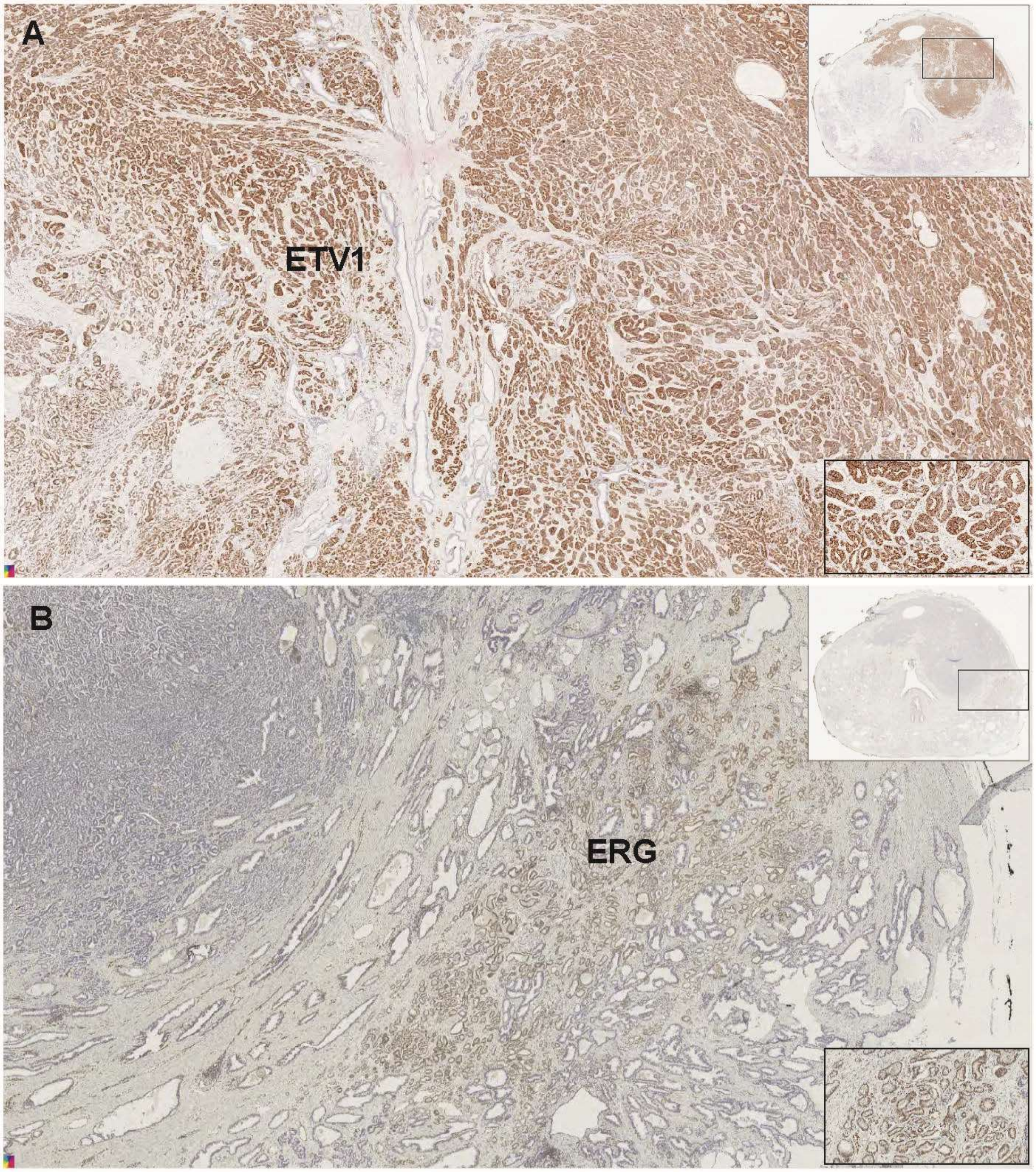


**Figure S2** : Radical prostatectomy tisse from a Caucasian American patient with prostatic adeno-carcinoma with Gleason score 3+4=7 (Grade Group 2) with extraprostatic extension or microscopic invasion of bladder neck, (pT3a). Large nodule occupies most of the right lobe in apex and mid region and extends anteriorly into anterior midline, left anterior and anterolateral region (apex and mid regions). Gleason 3=85%; Gleason 4=15% (fused and poorly formed glands) with tumor volume of 30%. A) Whole-mount tissue section stained for ETV1 and ETV4 show postive tumor foci for ETV1. B) Consecutive section stained with ERG and SPINK1 show ERG positive on the adjacent tumor foci. Inset at the top and bottom right show the wholemount and zoom in view, respectively.

Zhao et al., Figure S3

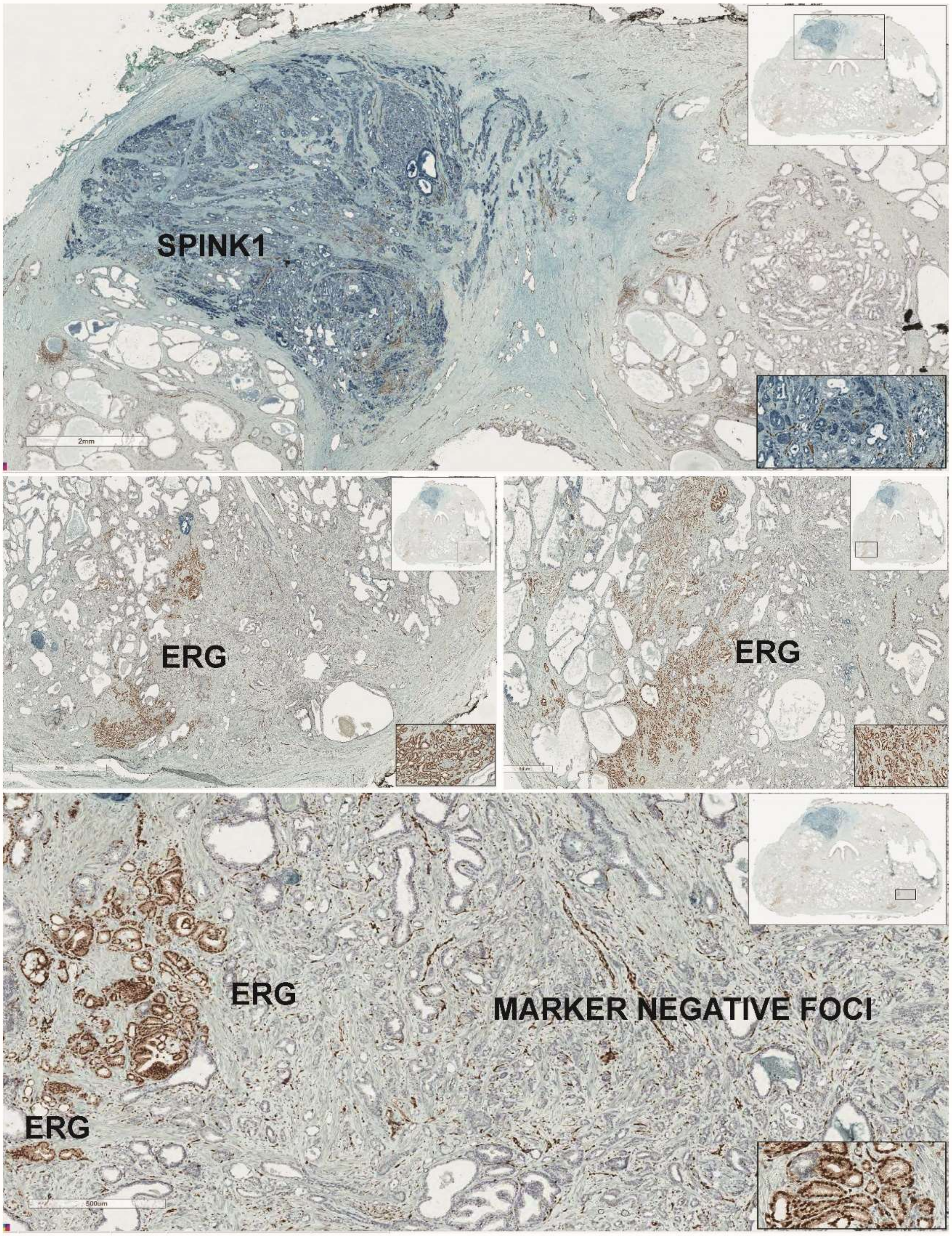


**Figure S3:** Wholemount tissue from an African Amerian patient positive for ERG and SPINK1 in multiple different tumor foci showing the extent of inter tumor heterogeneity. Inset at the top and bottom right show the wholemount and zoom in view respectively.

Zhao et al., Figure S4

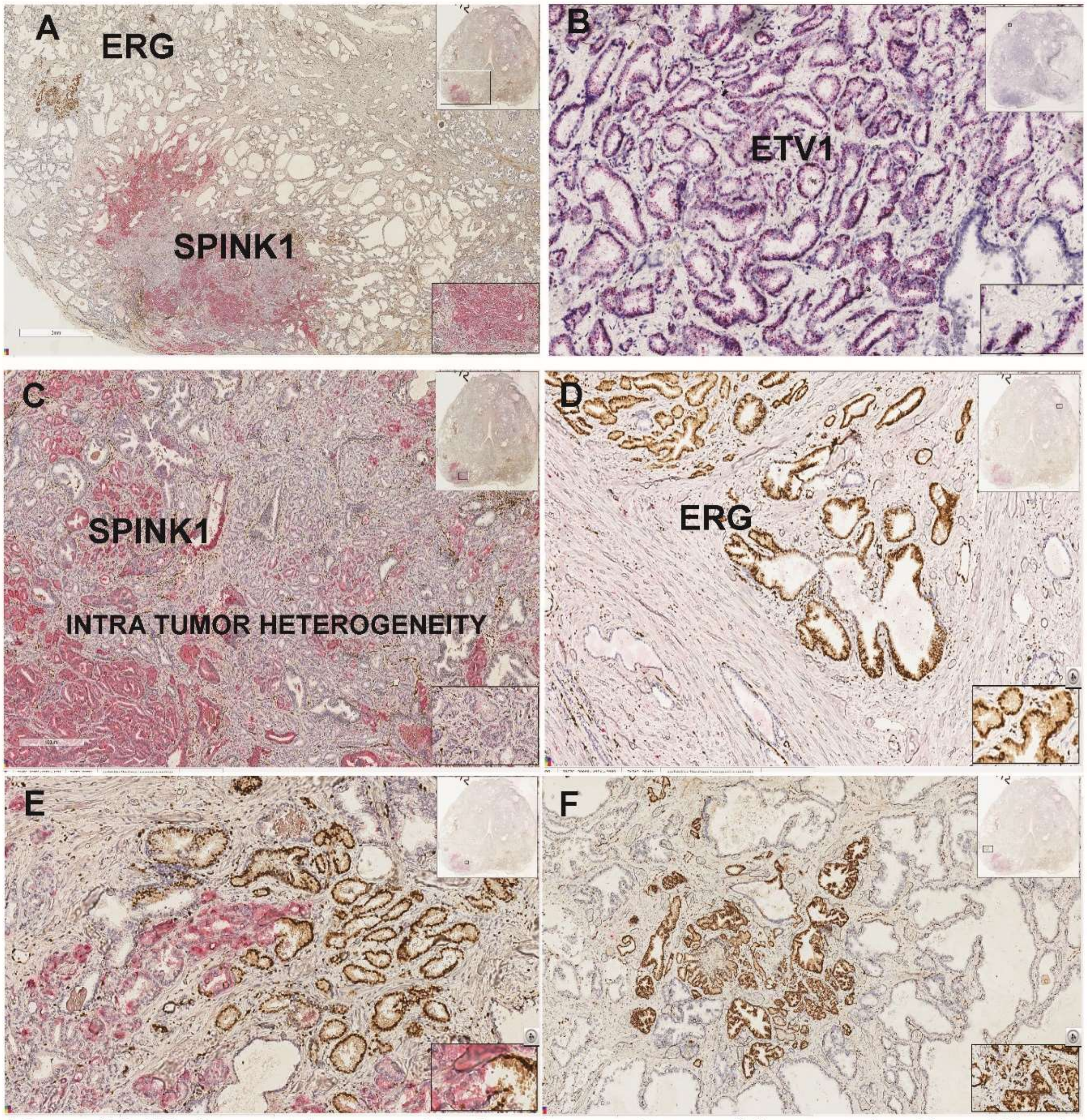


**Figure S4**: Wholemount tissue positive for adenocarcinoma of the prostate with multifocal cancer. Gleason grade 3+4=7 positive for ERG, SPINK1 and ETV1 in independent tumor foci. The tumor is organ confined with bilateral involvement. Adenocarcinoma extensively and multifocally involves bilateral anterior and right posterior at the apex, bilateral anterior and posterior at the mid, right anterior at the base of the gland. Gleason pattern 4 composes about 20% of the umor with tumor volume about 15%. ERG and SPINK1 multiple different foci. A) ERG and SPINK1 in two independent tumor foci. B) ETV1 postive tumor. C) Intra tumor heteroteneity in SPINK1 expression. D) Isolated HGPIN positive for ERG. E) Immediately adjacent tumor foci positive for ERG and SPINK1 in a mutually exclusive pattern. F) ERG positive tumor next to SPINK1 positive tumor foci. Inset at the top and bottom right shows the wholemount view with tumor location (rectangle box) and zoom in view of the tumor, respectively,

Zhao et al., Figure S5

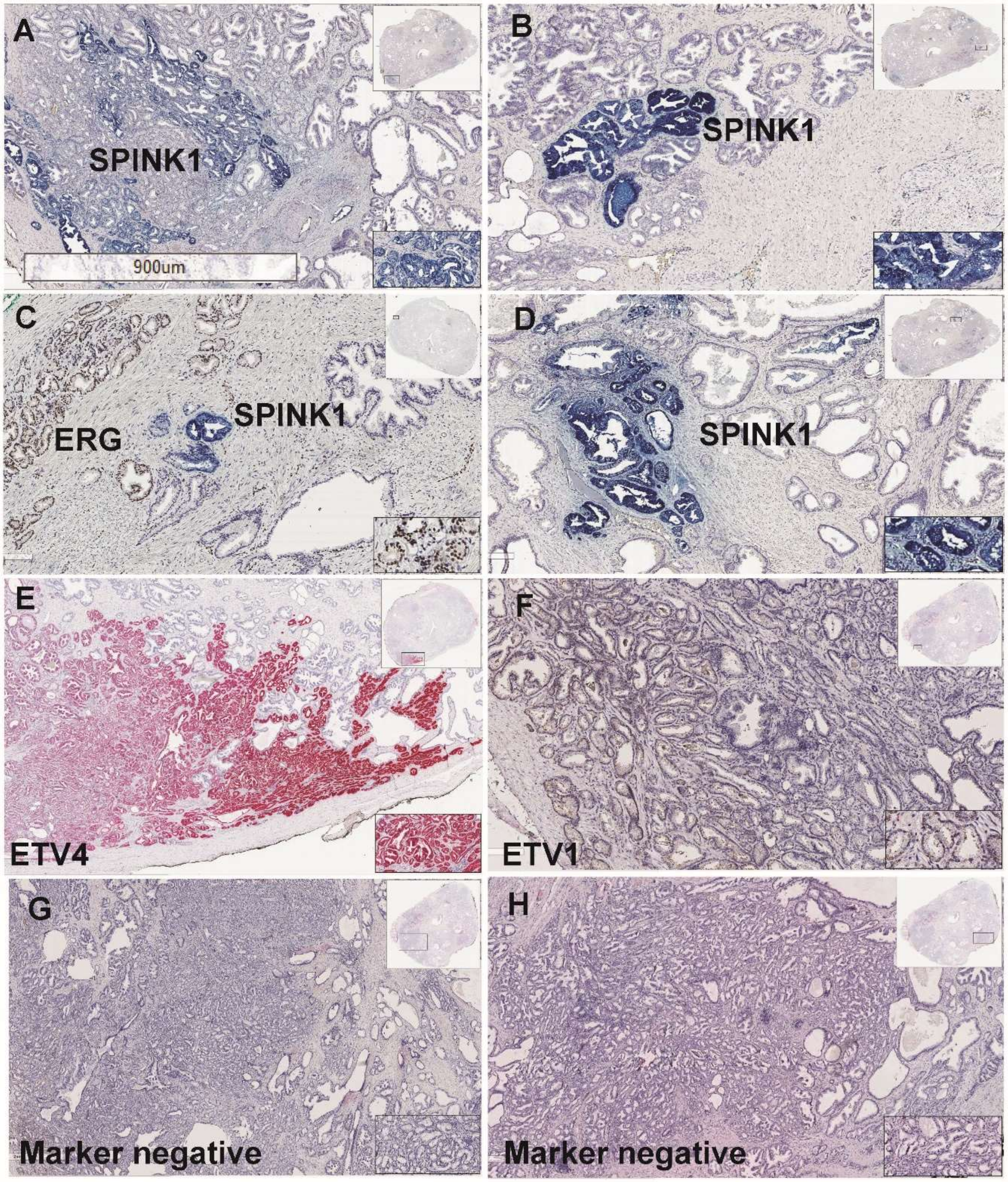


**Figure S5**: Wholemount tissue from a Caucasian American patient positive for ERG, SPINK1, ETV1 and ETV4. Organ confined pT2 stage cancer wiht large nodule in the left posterolateral/lateral region (apex to mid) with Grade Group 2 (Gleason score 3+4=7), 11-20% of tumor with Gleason 4 pattern and tumor at Left anterior apex with Grade Group 1. Multiple tumor foci are positive for SPINK1 (A-D). A tumor with hihg-grade prostatic intraepithelial neoplsia (HGPIN) is identified.All the four markers are present in a mutually exclusive pattern. ETV4 (E) and ETV1 (F) foci are located on two different tissue blocks. Tumors negative for all four markers (G &H).

Zhao et al., Figure S6

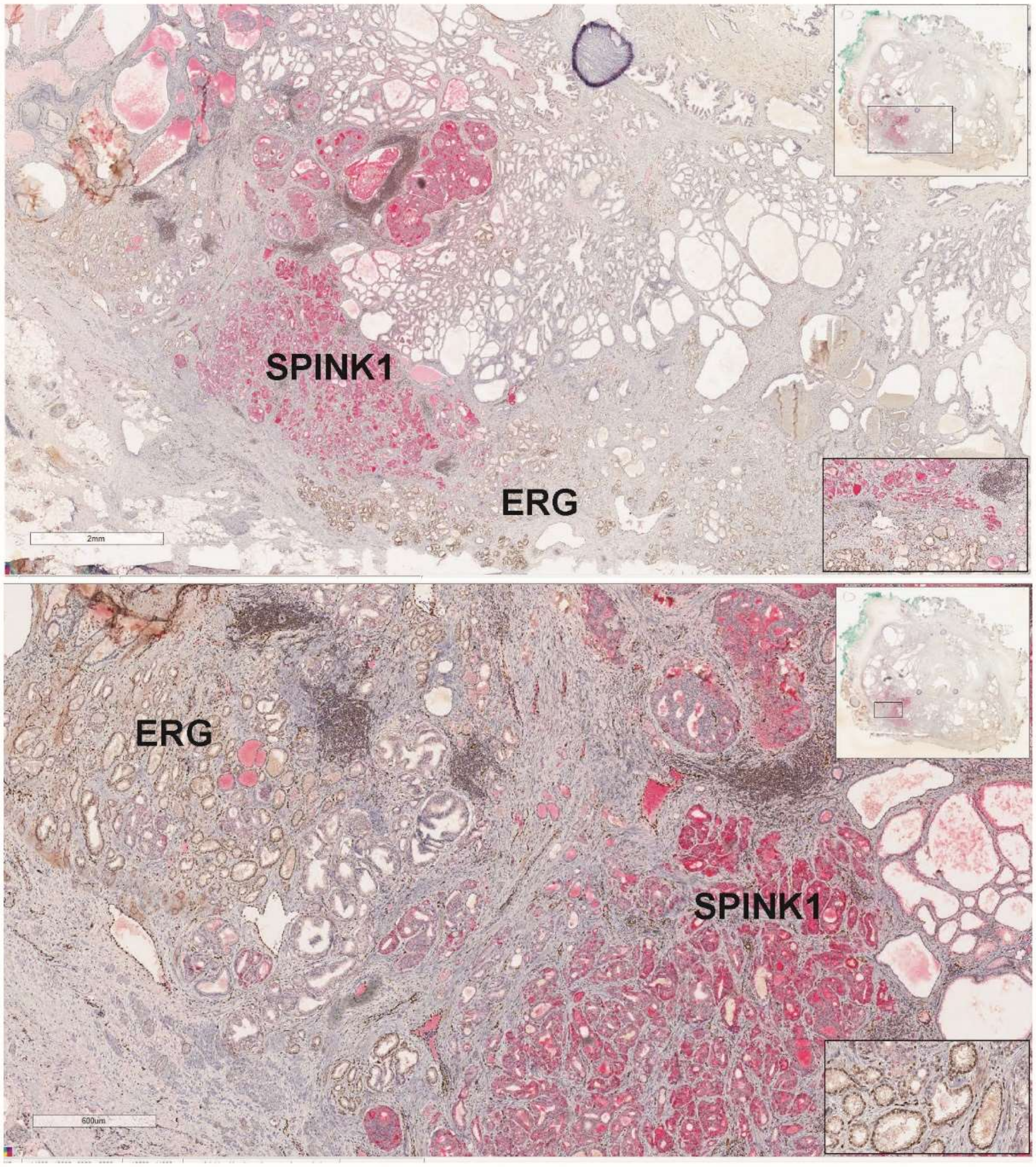


**Figure S6**: Wholemount tissue positive for ERG and SPINK1 in adjacent tumor foci in a mutually exclusive pattern. Prostate adenocarcinoma with Gleason score 7 93+4=7), organ confined with bilateral involvement (pT2c). Dominant nodule located right posterior/posterolateral region (apex to base) with a size of 3.6X1.7X1cm. Gleason 3=60%; Gleason 4=40%. Additional nodules at left posterolateral region (mid to base) with Gleason score 6(3+3=6), bilateral anterior/anterolateral (mid) with Gleason score 6 (3+3=6). The extent of inter tumor heterogeneity and mutually exclusive expression in independent tumor foci with varying morphological and Gleasonpattern indicates the extent of tumor molecular heterogeneity in prostate cancer. Inset at top and bottom right show tumor location in wholemount tissue and zoom in view of tumor positive for markers, respectively.

Zhao et al., Figure S7

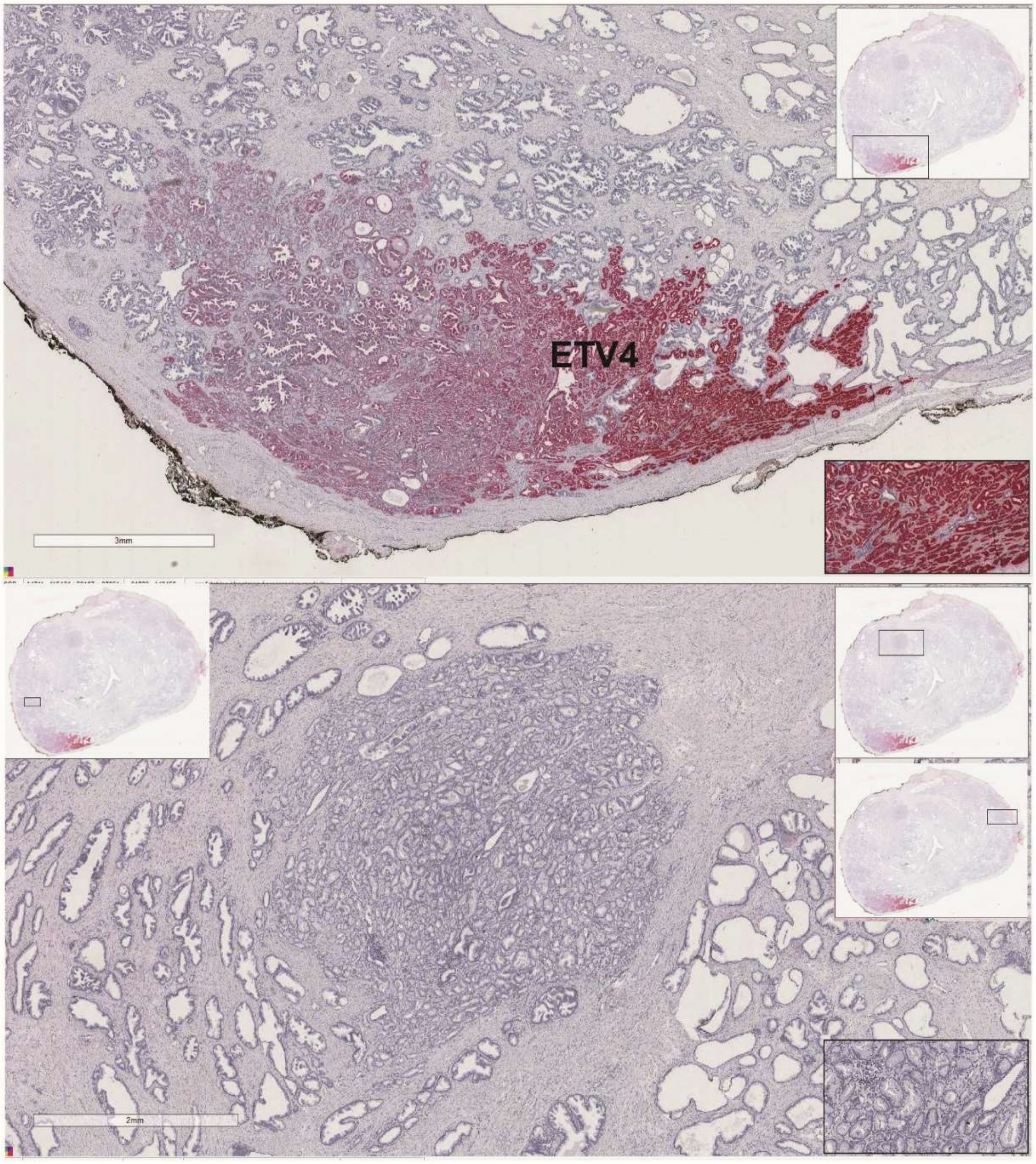


**Figure S7**: Whole mount tissue showing positive staining in one tumor foci and many other foci are negative indicationg the extent of inter tumorheterogeneity and independent clonal origin of tumor with distinct driver molecualr aberration. A tumor positive for ETV4 (red stain) with varying level of expression and four other tumor foci are negative both ETV1and ETV4 tested on this tissue.

Zhao et al., Figure S8

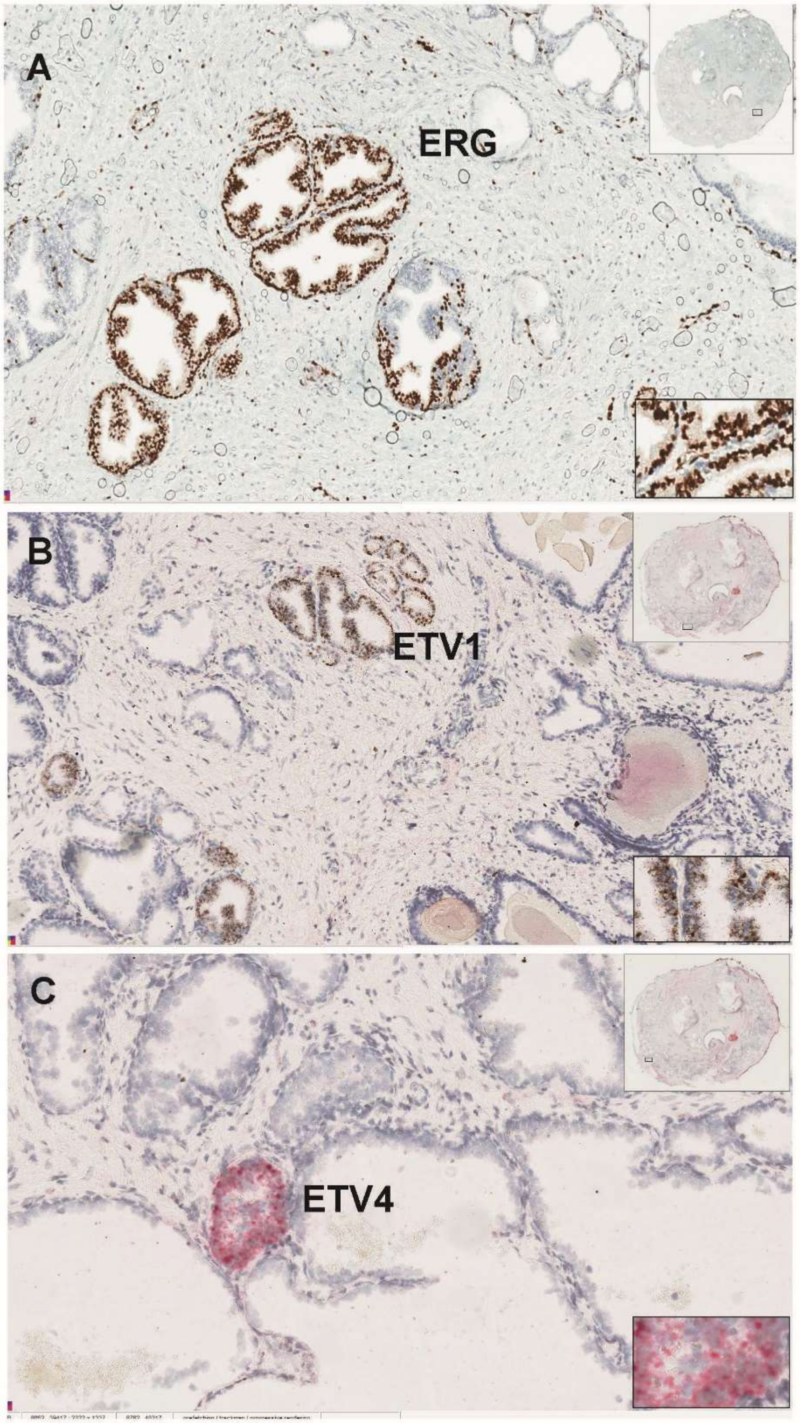


**Figure S8**:Wholemount tissue positive for ERG (A), ETV1 (B) and ETV4 (C) expression in three disticnt tumor foci. Prostate adenocarcinoma with Gleason score 3+4=7 (Grade group 2). Confined to the prostate, bilateral (pT2c). Gleason grade pattern 3=95%; pattern 4=5% (fused glands). Tumor volume 6%. located at bilateral anterior mid, left greater than right. Markers are positive in small isolated foci at different locations of the prostate. Insets at the top and bottom right show wholemount view and xoom in view of the tumors.
